# Genetic architecture of high-dimensional liver radiomic phenotypes and their role in common metabolic diseases

**DOI:** 10.64898/2026.05.19.26353617

**Authors:** Haodong Tian, Meghana Kamineni, Buu Truong, Vineet Raghu, Jacqueline S. Dron, Whitney Hornsby, Satoshi Koyama, Zhi Yu, Pradeep Natarajan

## Abstract

The liver plays a central role in systemic metabolism, yet large-scale genetic studies of quantitative liver imaging phenotypes remain limited. Here, we applied deep learning-based segmentation and radiomics extraction to derive 200 well-defined liver MRI features across multiple categories and imaging contrasts in 43,176 UK Biobank participants. Association analyses revealed steatosis-independent radiomic signals predicting incident chronic liver disease beyond conventional risk factors. We conducted genome-wide association studies in 37,725 individuals and identified multiple heritable liver MRI features; joint genetic structure and pleiotropy analyses demonstrated that these radiomic traits capture complex genetic architecture beyond the extent of hepatic steatosis. These MRI features showed widespread genetic overlap with plasma proteins, metabolites, and cardiometabolic traits through shared genetic loci and genetic correlations independent of adiposity. We identified putative causal links between liver MRI traits and cardiometabolic and liver-related outcomes, as well as evidence for pathway-specific imaging biomarkers to track activity of hepatically-influenced therapeutics.

## Introduction

The liver is central to systemic metabolic homeostasis, and hepatic dysfunction underlies a spectrum of common cardiometabolic diseases, including coronary artery disease, type 2 diabetes, and metabolic dysfunction-associated steatotic liver disease (MASLD). Magnetic resonance imaging (MRI) provides non-invasive, quantitative characterization of hepatic tissue at the population scale, and both epidemiological and genome-wide association studies (GWAS) of clinically-recognized MRI-derived liver phenotypes have yielded important insights into hepatic biology [1]. In particular, studies of proton density fat fraction (PDFF) have identified numerous associated loci, including well-established hepatic steatosis genes such as *PNPLA3* [2] and *TM6SF2* [3], and have systematically characterized their associations with metabolic traits and liver-related outcomes [4,5,6]. However, PDFF captures only a single summary of liver pathology. Abdominal MRI encodes richer biological information beyond global fat content, including spatial heterogeneity and tissue texture that may reflect processes such as fibrosis, inflammation, and architectural remodeling [7]. Radiomics provides a systematic framework for quantifying these high-dimensional imaging features across multiple contrasts, which may reflect underlying tissue biology [8,9]. Yet population-scale diverse liver MRI radiomic phenotypes remain scarce, leaving the genetic architecture of these traits largely unexplored, and it is unclear whether they capture heritable biological variation beyond conventional fat-based measures or reveal liver-specific genetic signals and causal pathways.

The increasing availability of large-scale population imaging and advances in automated image analysis now enable systematic genetic investigation of such high-dimensional liver phenotypes at scale. We applied deep learning-based segmentation and feature extraction to derive high-dimensional, well-defined liver MRI radiomic phenotypes and characterized their clinical relevance through epidemiological analyses. We performed GWAS to characterize their genetic architecture and examined their joint genetic structure relative to conventional liver PDFF. We evaluated their genetic overlap with circulating proteins, metabolites, cardiometabolic traits, and liver-related outcomes independent of obesity. We further investigated the causal connection of MRI traits to other clinical traits of interest and assessed their potential as pathway-specific imaging biomarkers of drug target genes. Figure 1 provides an overview of the study design and analyses.

**Fig 1.**
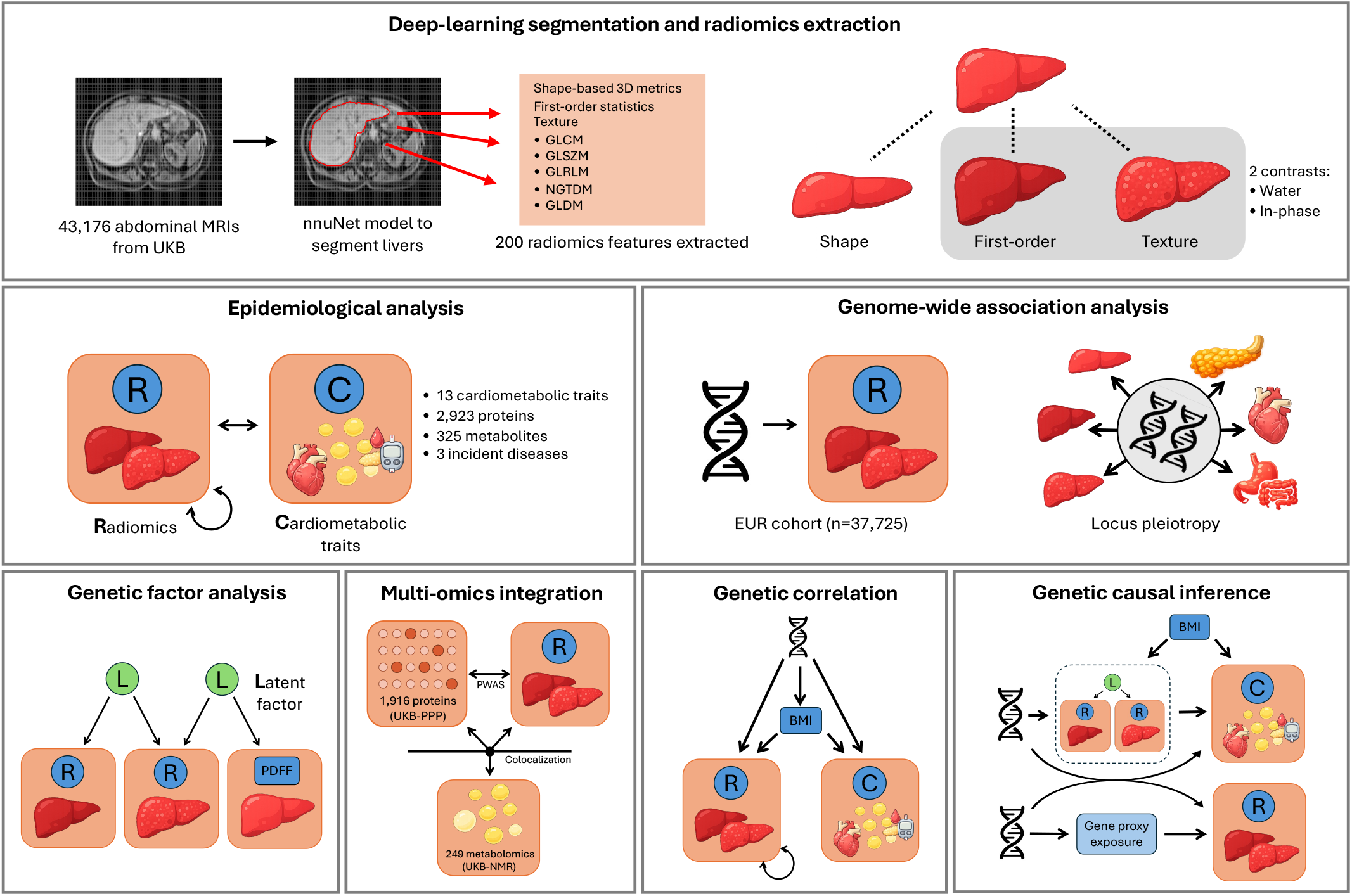
Overview of the study design and analytical framework. (Top) High-dimensional and well-defined liver MRI phenotypes were derived from 43,176 abdominal MRIs from UK Biobank using a deep-learning–based nnUNet segmentation model, yielding 200 radiomics features spanning shape-based 3D metrics, first-order statistics, and texture features (GLCM, GLSZM, GLRLM, NGTDM, and GlDM). (Middle) Phenotypic association analyses were conducted between radiomic traits and cardiometabolic traits, circulating proteins, NMR metabolites, and incident diseases, followed by hypothesis-free genome-wide association studies (GWAS) in a European cohort (n=37,725) to characterize the genetic architecture of liver MRI phenotypes, including locus pleiotropy across internal and external traits. (Bottom) Post-GWAS analyses comprised four complementary frameworks: genetic factor analysis to identify shared latent genetic structure across radiomic traits and liver PDFF; multi-omics integration linking radiomic traits to circulating proteins and NMR metabolites via proteome-wide association study (PWAS) and colocalization; genetic correlation to assess relationships between radiomic traits and external traits adjusting for BMI; and genetic causal inference via Mendelian randomization (MR) to evaluate putative causal relationships between radiomic traits and cardiometabolic outcomes, including drugtarget cis-MR to identify radiomic traits as pathway-specific imaging biomarkers.

## Results

### Deep learning-derived liver radiomics

Liver-specific images were extracted from the first abdominal MRI visit (commencing 2014) for 43,176 UK Biobank (UKB) participants with valid scans using deep learning segmentation. Radiomic features were subsequently derived using PyRadiomics (v3.1.0) [10] and organized into three main categories across two imaging contrasts (in-phase and water).

▪ **Shape features** were computed from the liver surface mesh independent of voxel intensity, using a marching cubes algorithm [11], and include interpretable morphological descriptors such as volume, axis lengths, and surface area (14 metrics).
▪ **First-order features** summarize the distribution of voxel intensities within the liver region of interest, capturing magnitude, uniformity, asymmetry, and standard statistical descriptors such as mean, median, and range (18 metrics, each per contrast).
▪ **Texture features** encode spatial patterns of gray-level intensity and are derived from five well-defined matrix-based representations: the gray level co-occurrence matrix (GLCM; 24 metrics), gray level size zone matrix (GLSZM; 16 metrics), gray level run length matrix (GLRLM; 16 metrics), neighboring gray tone difference matrix (NGTDM; 5 metrics), and gray level dependence matrix (GLDM; 14 metrics), each per contrast.

As shape features are contrast-independent, the final feature set comprises 200 liver MRI features per individual (14 shape + 93 x 2 contrast-specific features), spanning 3 main categories and 2 contrasts. Of the 43,176 participants, 96.6% reported White ancestry (n = 41,727), the mean (standard deviation) age was 64.06 (7.67), and 51.8% (n = 22,360) were female. For downstream GWAS, 37,725 individuals of European British ancestry were retained after additional genetic quality control. The imaging subcohort exhibited a mild healthy volunteer profile relative to the full UKB baseline population, with higher educational attainment and lower smoking rates (Supplementary Table S3).

Liver MRI features showed heavily skewed distributions (Figure 2a), with 72 of 200 features exceeding the absolute skewness of liver fat PDFF (|skewness| > 2.819), particularly among texture-based features. To mitigate the influence of extreme values while preserving interpretability, all features were winsorized prior to epidemiological analyses, reducing skewness to a range of -0.705 to 1.053 (Methods). After pruning features with pairwise Pearson |r| > 0.9, 84 features were retained for downstream analyses (11 shape, 38 water-based, and 35 in-phase non-shape features). PCA of the 84 retained features revealed a low-dimensional structure with gradual eigenvalue decay following the first component (Figure 2b). Effective dimensionality was lowest for texture features (7 of 59 PCs; 11.9%) and highest for first-order features (6 of 14 PCs; 42.9%), indicating that texture features carry substantial shared information and are summarized by fewer underlying dimensions. Notably, the newly derived MRI features recapitulate established liver MRI phenotypes; for example, *shape_SurfaceArea* was strongly correlated with UKB liver volume (Field ID 21080; Figure 2c), and a simple linear combination of *firstorder_Mean* from water and in-phase MRI closely approximated liver fat (Field ID 21088; Figure 2d).

**Fig 2.**
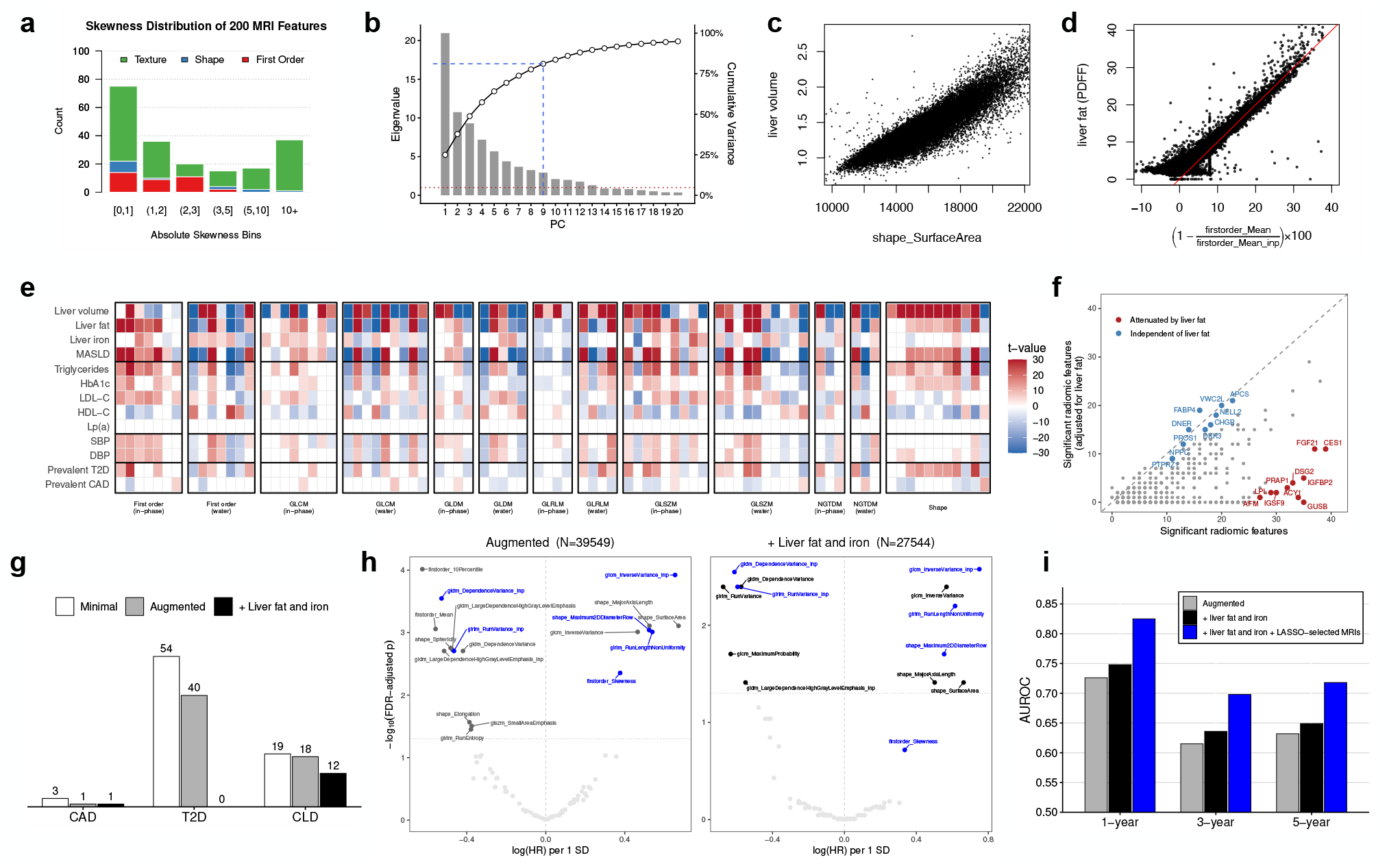
Epidemiological characterization of liver MRI features. (a) Distribution of absolute skewness across the 200 liver MRI features, stratified by radiomic category. (b) Principal component analysis of the 84 pruned MRI features. Bars denote eigenvalues for the first 20 principal components, and the overlaid line shows cumulative explained variance; dashed lines indicate the number of components required to explain approximately 80% of total variance. (c) Relationship between liver volume and a representative shape feature *(shape_SurfaceArea)*. (d) Relationship between liver fat (PDFF) and a first-order intensity contrast derived from water and in-phase images. (e) Heatmap of phenotypic associations between the 84 pruned MRI features and selected hepatic, metabolic, lipid, and cardiometabolic traits. Associations were estimated after adjustment for age, sex, and BMI; coloured tiles indicate statistically significant associations after global Bonferroni correction, with colour intensity representing the t value. MRI features are grouped by radiomic class and image contrast. MASLD, metabolic dysfunction-associated steatotic liver disease; Lp(a), lipoprotein(a); T2D, type 2 diabetes; CAD, coronary artery disease. (f) Comparison of MRI-protein association profiles before and after additional adjustment for liver fat. Each point represents one circulating protein, with the x axis showing the number of significantly associated MRI features in the base model and the y axis showing the number remaining after liver fat adjustment; selected labels highlight proteins with association profiles that were largely preserved or substantially attenuated. (g) Number of liver MRI features associated with incident chronic liver disease (CLD), coronary artery disease (CAD), and type 2 diabetes (T2D) across sequential Cox models. (h) Volcano plots for CLD in the augmented model and after further adjustment for liver fat and liver iron, with labels highlighting features retained in the multivariable LASSO model. (i) Time-dependent AUROC for prediction of incident CLD at 1, 3, and 5 years under progressively extended models, including the augmented model alone, the augmented model plus liver fat and liver iron, and the augmented model plus liver fat, liver iron, and LASSO-selected MRI features.

Phenotypic correlations between liver radiomic features and cardiometabolic traits revealed pervasive associations across MRI categories (Figure 2e). Most features showed concordant associations with liver fat, triglycerides, and MASLD, and inverse associations with HDL-C, consistent with hepatic lipid accumulation. Results were robust to replacing BMI with waist-to-hip ratio (WHR) as the adiposity covariate (Supplementary Figure S1). Proteome- and metabolome-wide analyses against 2,923 plasma proteins and 325 nuclear magnetic resonance (NMR) metabolites confirmed widespread associations (Supplementary Figure S2). After additional adjustment for liver PDFF, associations with canonical steatosis markers (e.g., FGF21, CES1, LPL) were substantially attenuated, whereas fibrosis- and remodelling-related proteins (e.g., APCS, DKK3, FABP4) were largely preserved (Figure 2f), reinforcing the distinction between fat-driven and fat-independent radiomic signals. To assess the clinical relevance of liver radiomic features, we examined their associations with three incident diseases using Cox proportional hazards models under progressively stringent covariate adjustment (minimal, augmented with disease-specific risk factors, and further adjusted for liver fat and iron). Radiomic features showed widespread associations with incident type 2 diabetes (T2D), but these were entirely attenuated after adjustment for liver fat and iron (Figure 2g), indicating a hepatic fat-driven signal. Associations with incident coronary artery disease (CAD) were weak throughout. In contrast, 12 radiomic features remained significantly associated with incident chronic liver disease (CLD) after full adjustment, including liver fat and iron, predominantly texture features, and LASSO variable selection under the most stringent model identified six of these as jointly independent predictors of incident CLD (Figure 2h). Addition of these LASSO-selected features to the clinical model improved 5-year AUROC for incident CLD from 0.649 to 0.718 (Figure 2i).

To complement these epidemiological findings with an unbiased genetic perspective, we performed genome-wide association analyses of all liver radiomic features.

### Genome-wide association results

We conducted GWAS on 200 liver MRI features in 37,725 individuals of European British ancestry from the UK Biobank. All MRI features were inverse-normal transformed prior to analysis to mitigate the influence of heavily skewed phenotypic distributions. LDSC intercepts (range: 0.99–1.03) indicated minimal confounding from population stratification or cryptic relatedness, while mild genomic inflation (λGC range: 1.02–1.15) was observed, likely due to the polygenic contributions from traits such as obesity (Supplementary Figure S4). To reduce redundancy, we retained 59 MRI features with pairwise genetic correlations |r_g| < 0.9 for downstream analyses, comprising 12 first-order, 7 shape, and 40 texture features, and 27 in-phase and 32 water-based features. The eigen-spectrum of the genetic correlation matrix across these 59 features showed a moderate and balanced eigenvalue decay (Figure 3a), with the leading six dimensions collectively explaining 80% of total genetic variance, indicating a highly structured but multidimensional genetic architecture not dominated by any single factor.

**Fig 3.**
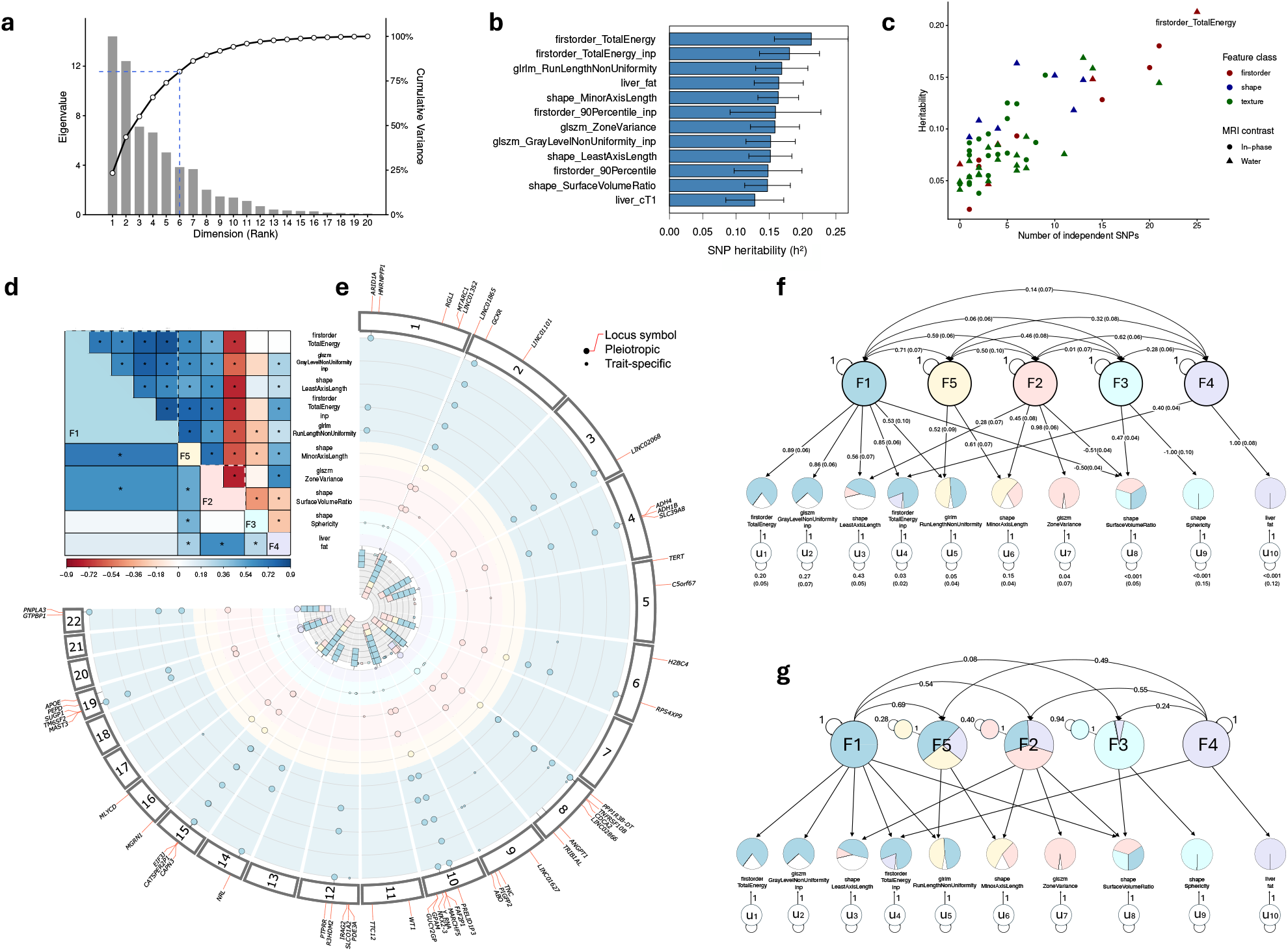
Genetic architecture of liver MRI features. (a) Eigendecomposition of the genetic correlation matrix across 59 retained liver MRI features. (b) SNP-heritability estimates for the 10 lead MRI features, together with liver fat (PDFF) and liver cT 1. (c) Relationship between SNP-heritability and the number of genome-wide significant independent SNPs (P < 5×10^−8^) across MRI categories and imaging contrasts. (d) Heatmap of pairwise genetic correlations among the 10 lead mRi features; clustering is based on the Genomic sEm factor structure results (see panel f), with each MRI feature assigned to the latent factor on which it exhibited the largest loading. (e) Internal pleiotropy analysis results for the 10 lead MRI features. Each dot corresponds to a trait-associated locus. Larger dots highlight a pleiotropic association. The internal bar plot represents the number of associated traits for each pleiotropic locus. (f) Path diagram of the five-factor Genomic SEM model fitted to the 10 lead MRI features. Pie plots of the MRI features indicate the proportion of genetic variance explained by each latent factor (F1—F5); inter-factor correlations are omitted in the pie plot for visual clarity. All effect sizes are standardized. (g) Path diagram of the hierarchical model fitted to the five latent factors. Pie plots for factors indicate the proportion of variance explained by the two higher-order factors, represented by the two independent factors F1 and F4.

Single-nucleotide polymorphism (SNP)-based heritability estimates across the 59 MRI features ranged from 0.023 to 0.213 (Supplementary Table S4). The 10 most heritable features showed heritabilities comparable to those of liver fat and liver iron-corrected T1 (cT1) (Figure 3b), with the two highest being first-order total energy derived from the in-phase and water images. This pattern was not simply driven by overlap with conventional liver MRI traits: for example, *firstorder_TotalEnergy* showed essentially no genetic correlation with liver fat and liver cT1, and the most heritable MRI features overall displayed heterogeneous relationships with both traits (Supplementary Table S6). Intensity-based features directly reflect hepatocellular composition, including lipid, water, and iron content. SNP-heritability was positively associated with the number of genome-wide significant independent variants (Figure 3c), and no systematic clustering by feature category or imaging contrast was observed (Supplementary Figure S5), suggesting that genetic informativeness is not restricted to any particular MRI modality or feature type.

We identified 327 genome-wide significant loci (P < 5×10^−8^) across the 59 MRI features, and 57 loci for the 10 lead features, of which 21 exhibited cross-feature pleiotropy (Figure 3e; Supplementary Table S8). Loci at *GCKR* and *CATSPER2P1 /TP53BP1* were associated with all lead features except *shape_Sphericity* and liver fat, and are annotated to fatty acid and triglyceride metabolism in the EMBL-EBI GWAS Catalog. Four of the ten liver fat loci *(PNPLA3, TM6SF2, APOE*, and *TRIB1)* showed pleiotropy with other lead MRI features, while 47 lead-MRI loci were not identified in prior liver fat GWAS [6]. To further characterize the liver fat loci, we compared the effects of *PNPLA3* (rs738409, p.Ile148Met), *TM6SF2* (rs58542926, p.Glu167Lys), and *APOE* (rs429358, p.Cys130Arg) across all 59 MRI features (Supplementary Figure S6), where *PNPLA3* and *TM6SF2* variants showed highly concordant effect profiles, consistent with their shared downstream consequence of hepatic triglyceride accumulation, whereas *APOE* variants effects were more dispersed, reflecting its distinct mechanism of action through lipoprotein metabolism. The internal pleiotropy patterns were broadly concordant with the genetic correlation structure, with pleiotropic effects concentrated within features sharing the same latent factor (Figure 3d–e). Among 19 novel lead-MRI variants across 15 loci not previously reported in the GWAS Catalog, more than half (10/15) were specific to shape-based features. Partitioned LDSC analysis revealed consistent enrichment of heritability in liver-specific annotations across all nine lead MRI features (2.91-to 5.64-fold), generally exceeding that of liver fat (3.44-fold), though enrichment was not conditionally significant after accounting for baseline-LD model annotations, suggesting that general regulatory features could largely underlie the observed liver-specific signal (Supplementary Table S11).

Replication was performed using a held-out UKB discovery/validation split (80%/20%; n = 30,227/7,564). Of 274 genome-wide significant variants identified in the discovery set, 120 (43.8%) and 166 (60.6%) replicated at Bonferroni-corrected and nominal thresholds, respectively, with 262 (95.6%) showing concordant effect directions. In the non-European UKB independent set (n = 3,895), 307 of 342 variants (89.8%) showed concordant directions, with replication rates of 38.6% (nominal) and 22.2% (Bonferroni-corrected). These replication rates are consistent with expectations given the respective sample sizes and the known tendency for initial GWAS discoveries to modestly overestimate effect sizes.

### Genetic structure between MRI features and liver fat

Liver MRI features may reflect multiple underlying biological pathways, each represented by distinct latent genetic factors. We selected nine lead MRI features with the highest LDSC-based heritability z-scores (>7.5), spanning shape, first-order, and texture categories, alongside liver fat, and applied Genomic SEM [12] to characterize their latent genetic structure (Methods). Features with similar genetic correlations tended to load onto shared factors, whereas those with weaker or distinct correlations were captured by separate factors, suggesting a heterogeneous genetic architecture across liver MRI phenotypes.

Among candidate models, a five-factor solution provided the best fit (AIC = 297.01, CFI = 0.998, SRMR = 0.052; Figure 3f; Supplementary Table S14-S15). Several MRI features are loaded by multiple factors simultaneously, reflecting the complexity of liver imaging phenotypes, and individual MRI features are rarely exclusive biomarkers of a latent factor. This raises caution for multivariate GWAS approaches (e.g., MTAG [13]) that assume a single common factor model, as different genetic variants may influence the same MRI feature through distinct biological pathways.

Factors F3 and F4 loaded exclusively onto *shape_Sphericity* and *liver_fat*, respectively, with near-zero residual genetic variance, indicating that these two factors can be fully captured by their respective MRI features. Notably, F4 (liver fat) cross-loaded with only one additional feature, *firstorder_TotalEnergy* (in-phase), likely reflecting the fat-sensitive nature of in-phase MRI acquisition. This highlights that liver fat is genetically distinct from most other derived MRI features, which are not co-loaded onto the same latent factor, suggesting that the broader MRI feature set captures latent hepatic pathways beyond what PDFF alone can reveal. Across all ten features, the median residual genetic variance unexplained by latent factors was 3.66%, supporting a largely shared and lowdimensional genetic structure; *shape_LeastAxisLength* was a notable exception, with 43% unexplained variance, indicating a more distinct genetic architecture. We additionally fitted a hierarchical two-factor model (CFI = 0.995, SRMR = 0.073), which indicated that the two higher-order factors were primarily explained by F1 and F4 (Figure 3g; Supplementary Table S16-S17). Together, these results validate the genetic information captured by our novel MRI features beyond liver fat, while demonstrating that their complex latent structure may render simple multi-trait frameworks probably ill-suited for liver imaging genetics.

### External pleiotropy analysis

To better investigate the external pleiotropy of liver MRI feature-associated variants, we leveraged PLATLAS [39], a large-scale pleiotropy platform comprising >1,000 traits from major global biobanks, and queried the 51 variants of the 9 lead MRI features (grouped into F1, F2, F3, F5 by Genomic SEM) across 1,119 traits in 18 categories using EUR-ancestry GWAMA summary statistics (Bonferroni-corrected threshold: P < 0.05/[1,119 × 51]; Methods). Among the 18 PLATLAS trait categories, the strongest external pleiotropy signals were observed in Endocrine/Metabolic, Circulatory System, and Digestive categories (Figure 4; Supplementary Table S9-S10), consistent with the established role of the liver in metabolic regulation, lipid homeostasis, and digestive function. Notably, variants associated with genetic factor F2 *(glszm_ZoneVariance, shape_SurfaceVolumeRatio)* exhibited a disproportionately higher number of pleiotropic associations compared to variants in other factor groups, including the two PDFF variants rs738408 *(PNPLA3)* and rs58542926 *(TM6SF2)* that show similar internal and external pleiotropy patterns. This suggests that F2-related liver imaging traits may capture broader systemic metabolic processes. In contrast, 21 of the 51 variants (41.2%) showed no detectable external pleiotropy in the PLATLAS dataset, which indicates that these signals are largely liver-specific and may provide insights into novel biological mechanisms not captured by existing disease GWAS.

**Fig 4.**
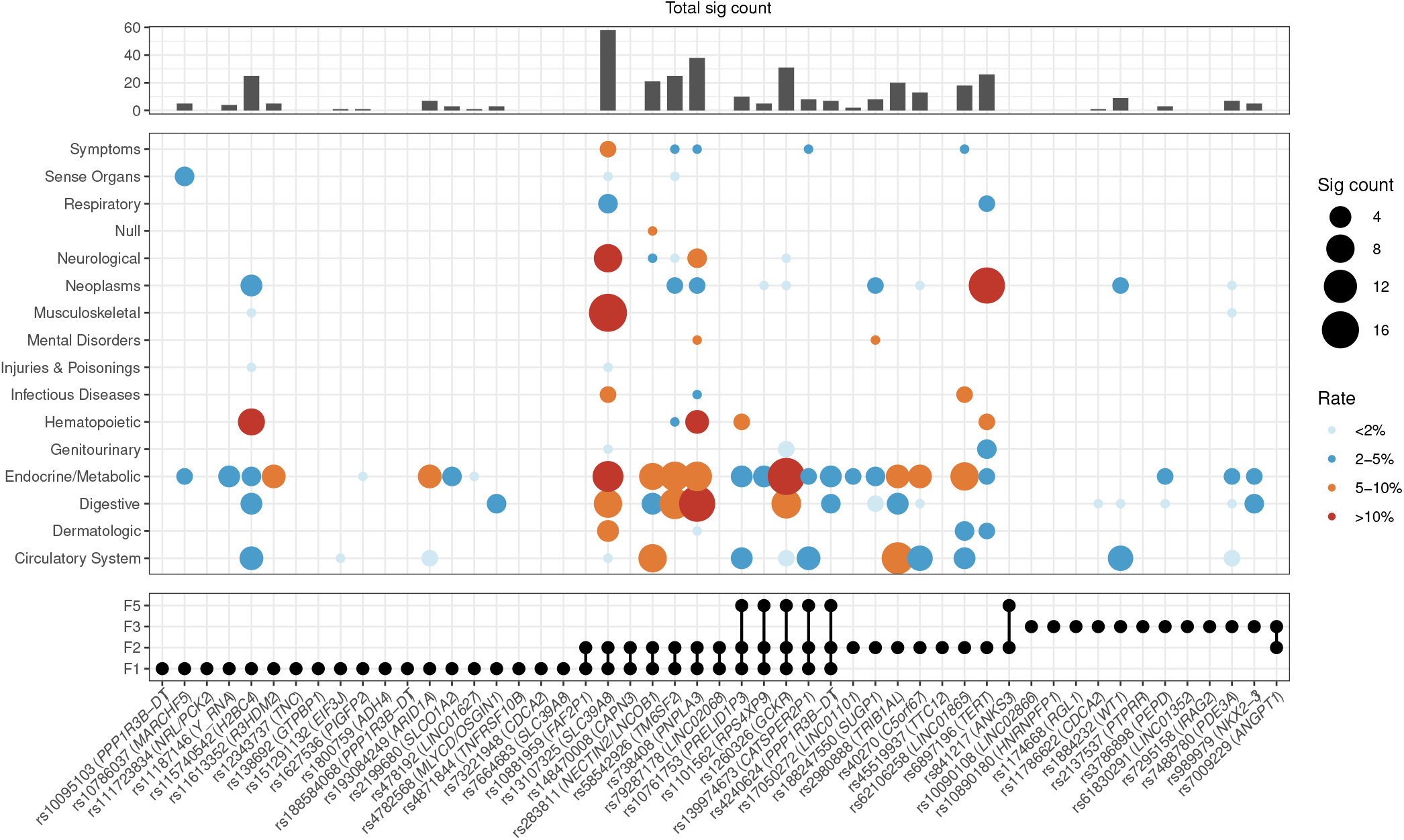
External pleiotropy of 51 liver MRI feature-associated variants across 18 PLATLAS categories (1119 traits). Each bubble represents the pleiotropy signal of a variant (x-axis) within a PLATLAS trait category (y-axis). Bubble size indicates the number of significantly associated traits (Bonferroni-corrected threshold: P < 0.05/[1,119 x 51]), and bubble color reflects the proportion of significant traits within each category (<2%, 2–5%, 5–10%, >10%). The bar chart (top) summarizes the total number of significant trait associations per variant across all categories. The dot plot (bottom) displays the internal pleiotropy pattern of each variant across the four Genomic SEM factors (F1, F2, F3, F5), with connected dots indicating variants with pleiotropic effects on multiple factors. Factor compositions are as follows — F1: *firstorder_TotalEnergy, firstorder_TotalEnergy_inp, glrlm_RunLengthNonUniformity, glszm_GrayLevelNonUniformity_inp, shape_LeastAxisLength*; F2: *glszm_ZoneVariance, shape_SurfaceVolumeRatio;* F3: *shape_Sphericity;* F5: *shape_MinorAxisLength*. Variants are ordered by factor group assignment. Traits not belonging to clearly defined categories are denoted as NULL in PLATLAS. Only variants with at least one significant association are represented by bubbles; variants with no detectable external pleiotropy appear on the x-axis without bubbles.

### Proteomic and metabolomic architecture of liver MRI signals

To systematically evaluate the relationship between circulating proteins and liver MRI radiomic features, we performed a proteome-wide association study (PWAS) with non-overlapping discovery and testing samples (Methods). Across 1,916 proteins from the UK Biobank Pharma Proteomics Project (UKB-PPP) and 59 liver MRI traits, PWAS identified 93 significant protein–MRI pairs at global FDR < 0.05, involving 28 proteins and 37 MRI features. These proteins spanned lipid/metabolic, fibrosis/extracellular matrix (ECM), and immune/inflammatory categories (Supplementary Figure S9). Among them, the loci indexed by NCAN and SERPINA1 showed the broadest patterns of association across MRI traits (Figure 5a). Colocalization analysis further showed that 47 of the 93 pairs (51%) were supported by shared genetic signals (PP.H4 > 0.8) (Supplementary Table S20). Notably, only one MRI feature remained colocalized with NCAN, suggesting that the broader PWAS signal at this locus may reflect the nearby *TM6SF2* region rather than circulating NCAN itself. APOE showed the broadest colocalization across MRI traits, whereas INHBB/INHBC and TNFSF10 each colocalized with multiple features from distinct radiomic domains. Overall, the colocalized architecture was concentrated in a subset of hub proteins rather than being uniformly distributed across all associations, and colocalized signals spanned first-order, texture, and shape-related traits (Figure 5b).

**Fig 5.**
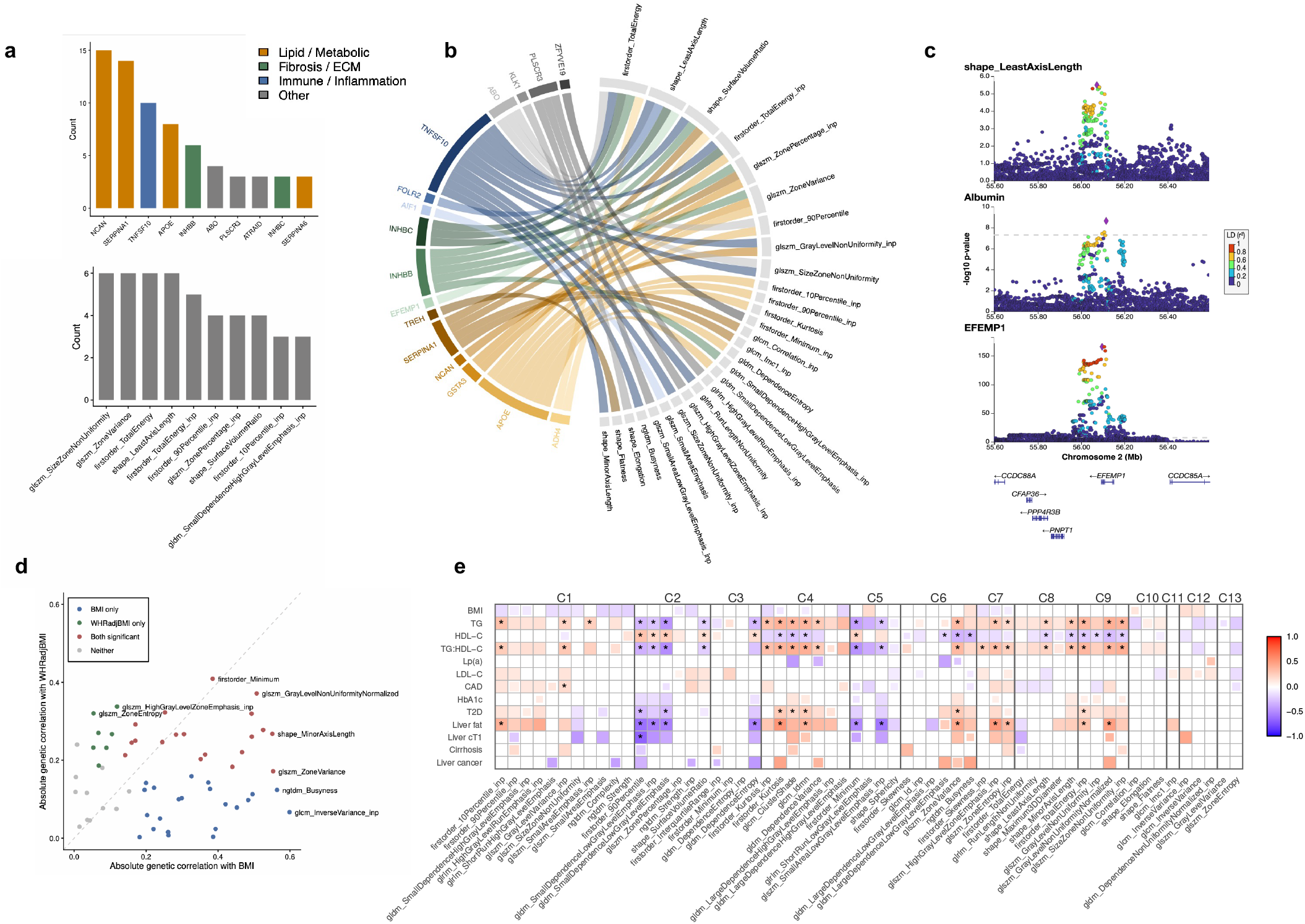
Genetic relationships of liver MRI radiomic features with circulating proteins, metabolites and related complex traits. (a) Summary of the top 10 proteins and top 10 MRI features with the largest numbers of significant PWAS signals across all 1,916 x 59 protein-MRI tests (global FDR < 0.05). Proteins are grouped into broad biological categories, including lipid/metabolic, fibrosis/extracellular matrix (ECM) and immune/inflammatory components. (b) Chord diagram showing protein-MRI pairs with evidence of colocalization (PP.H4 > 0.8) among the PwAS-significant associations. In total, 47 colocalized pairs were identified, involving 16 of 28 proteins and 28 of 37 MRI features represented in the PWAS-significant set. (c) Regional association plots at the *EFEMP1* locus for plasma EFEMP1, serum albumin (PP.H4 = 0.98) and the MRI feature shape_LeastAxisLength (PP.H4 = 0.82), illustrating a shared genetic signal across protein, metabolite and liver MRI traits. (d) Scatter plot of the 59 MRI features according to their genetic correlation estimates with BMI and WHRadjBMI. MRI features are grouped into four categories based on Bonferroni significance (P < 0.05/59) for genetic correlation with BMI only, WHRadjBMI only, both traits or neither trait. (e) Heatmap of genetic correlations between 59 BMI-adjusted (instance 0) liver MRI features and selected cardiometabolic traits. MRI features are grouped into 13 clusters (C1-C13) based on their internal genetic correlation structure. Nominally significant entries (P < 0.05) are lightly shaded; entries passing global FDR correction (P < 0.05) are fully colored; asterisks denote entries surviving global Bonferroni correction. TG, triglycerides; LDL-C, low-density lipoprotein cholesterol; HDL-C, high-density lipoprotein cholesterol; cT1, iron-corrected T1.

For the 16 colocalized loci, we further investigated associations with 249 metabolites measured in the UKB-NMR cohort (n = 434,646) [14], using a significance threshold of P < 0.05/(16 × 249) based on lead cis-pQTL associations. These loci showed marked heterogeneity in their metabolomic footprints, ranging from broad hub-like loci (for example, *APOE* and *ABO)* with extensive NMR associations to more selective loci (for example, *EFEMP1)* with little or no detectable metabolomic signal (Supplementary Table S21). At the *EFEMP1* locus, only one NMR trait, albumin, showed a significant association. Plasma EFEMP1 colocalized with both serum albumin (PP.H4 = 0.98) and hepatic *shape_LeastAxisLength* (PP.H4 = 0.82), supporting a shared causal variant across proteomic, metabolomic and imaging domains (Figure 5c), implicating ECM-mediated liver structural remodeling as a determinant of hepatic synthetic function.

### Genetic correlation analysis with cardiometabolic and liver-related traits

We investigated genetic correlations between 59 MRI features and external traits using cross-trait LDSC regression, encompassing 249 NMR metabolites and 13 specific cardiometabolic and liver-related traits. We first considered the 249 NMR metabolites and found pervasive genetic correlation signals (Supplementary Figure S11). Genetic correlations with BMI and WHRadjBMI revealed heterogeneous patterns across the 59 MRI features (Bonferroni threshold, P < 0.05/59; Figure 5d). The majority of features were genetically correlated with BMI alone (n = 22) or both traits (n = 19), highlighting overall adiposity as a pervasive source of genetic confounding. A smaller subset showed selective correlation with WHRadjBMI (n = 7), suggesting additional contributions from fat distribution. Given that BMI confounds many cardiometabolic traits of interest, we repeated GWAS with BMI-adjusted MRI phenotypes and reran the genetic correlation analyses. Adjusting for BMI measured at instance 0 yielded cleaner genetic correlation patterns than adjustment at instance 2 (Supplementary Figure S11, right), likely because earlier BMI better captures latent adiposity, while later BMI adjustment may introduce more severe collider bias. We therefore adopted BMI (instance 0)-adjusted results as primary. BMI-adjusted MRI features retained similar SNP-heritability to unadjusted features, and their genetic correlations with external traits were broadly proportional to unadjusted estimates (Supplementary Figure S12-S13, with attenuation being more pronounced for insulin resistance-related traits such as T2D and triglyceride to high-density lipoprotein cholesterol (TG:HDL-C) ratio.

After BMI adjustment, the strongest and most widespread genetic correlation signals (Bonferroni-corrected P < 0.05) were observed with TG, HDL-C, TG:HDL-C ratio, T2D, and liver fat (Figure 5e). Since TG:HDL-C ratio is strongly correlated with insulin resistance [15], and liver fat has a bidirectional relationship with insulin resistance, these results suggest that insulin resistance may represent a dominant shared pathway underlying the genetic correlations between liver MRI features and cardiometabolic traits. Despite this overarching pattern, individual MRI features showed trait-specific signals. For instance, *firstorder_Minimum_inp* (cluster C3) was significantly correlated with LDL-C only (global FDR P < 0.05), and several features in clusters C10–C13 (e.g., *glszm_ZoneEntropy)* correlated with lipid traits but not with liver-related phenotypes, highlighting genetic information beyond conventional liver MRI measures such as PDFF and cT1. Notably, *firstorder_Skewness* (cluster C6) showed a significant genetic correlation with cirrhosis (global FDR P < 0.05) but not with liver cT1, suggesting that some liver MRI features may capture cirrhosis-related structural variation beyond fibrosis-related information reflected by cT1.

These findings suggest that different liver MRI features capture distinct biological pathways, with insulin resistance likely accounting for the largest share of the observed genetic correlation signals. Consistent with this interpretation, metabolic traits more directly linked to insulin resistance, such as TG and T2D, showed broader and stronger genetic correlations with MRI features compared to Lp(a), LDL-C, and CAD (Supplementary Figure S14. For CAD specifically, most correlated MRI features (e.g. *ngtdm_Strength_inp* and *glszm_GrayLevelVariance_inp)* were also among the features genetically correlated with LDL-C in the same direction, suggesting that LDL-C may be the major mediator of the genetic correlation between these MRI features and CAD risk.

### Radiomics-wide Mendelian randomization

In light of the widespread genetic correlations identified, we investigated potential causal relationships between liver MRI features and cardiometabolic traits via Mendelian randomization (MR). Given the complex latent factor structure and confounding role of obesity indicated by the previous analysis, we considered multiple causal scenarios (Figure 6a) and adopted a comprehensive MR framework accounting for potential confounding from BMI (Methods). We aim to identify MRI features causally connected to each outcome, either through a direct effect (Scenario 1 ; Figure 6a) or as surrogates of a latent liver factor with downstream causal effects (Scenarios 2 &3; Figure 6a).

**Fig 6.**
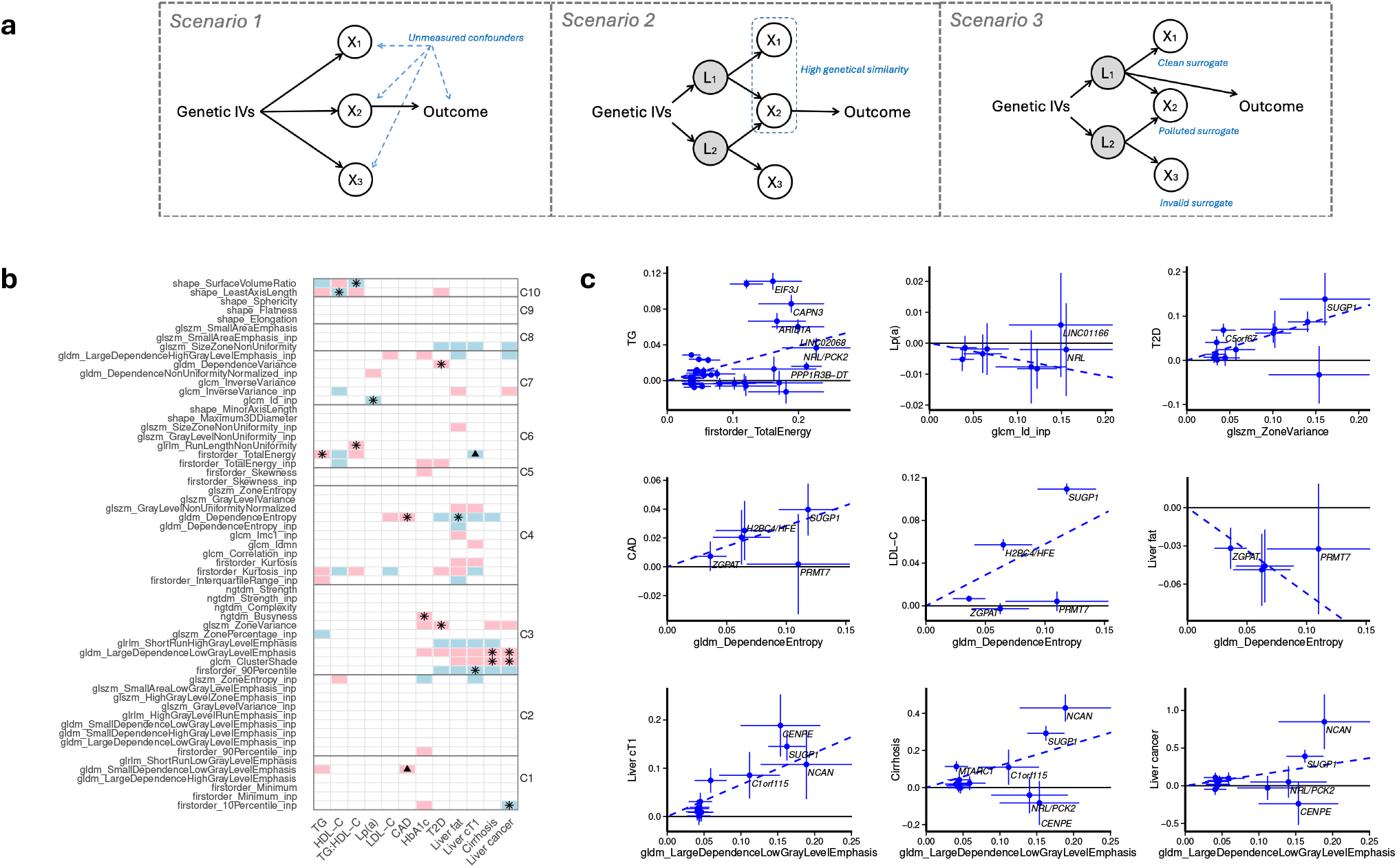
Mendelian randomization analysis of liver MRI features on cardiometabolic and liver-related outcomes. (a) Schematic illustration of possible causal scenarios linking liver MRI features to outcomes of interest. (b) Heatmap of highlighted MRI feature-outcome pairs with FDR P < 0.05 within each outcome. The 59 MRI features are grouped into 10 clusters according to the internal genetic similarity indicated by the joint genetic instruments. Red and blue entries denote positive and negative MR effect directions, respectively; stars and triangles indicate MRI features highlighted by MVMR-SuSiE. (c) MR scatter plots for selected MRI featureoutcome pairs, showing per-variant genetic association estimates with 95% confidence intervals and annotated nearest gene names. TG, triglycerides; LDL-C, low-density lipoprotein cholesterol; HDL-C, high-density lipoprotein cholesterol; cT1, iron-corrected T1.

MR identified multiple MRI features with potentially causal connections across all traits examined, with signal distributions varying by outcome (Figure 6b). To address potential redundancy arising from genetic correlations among MRI exposures, we applied MVMR-SuSiE, considering all highlighted MRI features jointly to select the key features (Methods). We found that most outcomes have only one key MRI feature, representing probably one latent causal pathway. For CAD, two MRI features instead emerged with distinct causal pathways: genetically predicted *gldm_DependenceEntropy* was associated with LDL-C (concordant direction), liver fat, and T2D (discordant direction), while the genetically predicted *gldm_SmallDependenceLowGrayLevelEmphasis* was instead associated with TG. Among the variants for *gldm_DependenceEntropy*, rs8107974 is an intronic variant in *SUGP1*, a gene implicated in cholesterol homeostasis through modulation of hepatic receptor splicing [16], and rs1800562 p.Cys282Tyr is a missense variant in *HFE*, which may promote atherosclerosis via iron-induced oxidative stress and disruption of hepatic lipid metabolism.

Relative to lipid traits, more causal signals were identified for liver-related outcomes (liver fat, cT1, cirrhosis, and liver cancer) and glycemic traits (T2D, HbA1c), a pattern corroborated by sensitivity analyses using MVMR-FA (Methods and Supplementary Note S2). Notably, several water-based MRI features, including *gldm_LargeDependenceLowGrayLevelEmphasis, glcm_ClusterShade*, and *firstorder_90Percentile*, showed consistent causal effect directions across liver fat, liver cT1, cirrhosis, and liver cancer. Given that liver cT1 reflects fibro-inflammatory liver injury [1], this pattern suggests that water-based features may capture tissue-level pathological changes beyond hepatic fat content, partly related to fibrosis-related remodeling, while potentially also encoding broader structural alterations underlying multiple liver outcomes.

### Drug-target cis-MR analysis

We investigated whether liver MRI features can serve as imaging biomarkers for specific drug-target gene pathways (illustrated by Figure 7a) on six therapeutically relevant genes with liver influence *(GLP1R, LPA, PCSK9, HMGCR, APOC3*, and *ANGPTL3)*. We applied several cis-MR analyses (MR-IVW, MR-PC-GMM, and MR-LD-IVW) with multiple data sources (GTEx, UKB-PPP, and GWAS), along with positive and negative controls (Methods).

**Fig 7.**
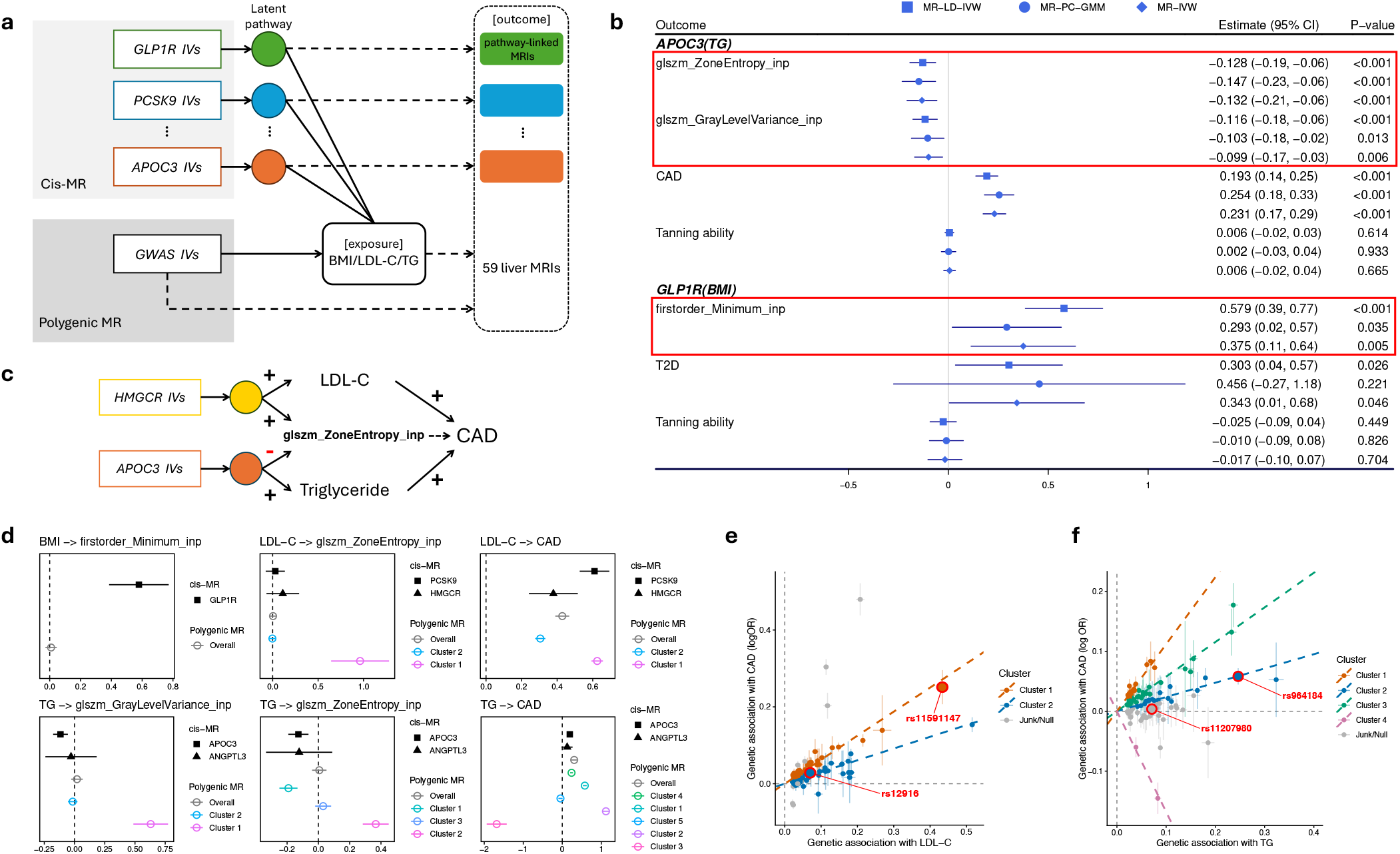
Drug-target cis-MR analysis of liver MRI features. (a) Schematic illustration of the analytical framework for identifying drug-target pathway-linked liver MRI biomarkers using cis-MR and polygenic MR. (b) cis-MR results (MR-IVW, MR-LD-IVW, and MR-PC-GMM) for highlighted gene-MRI feature pairs passing FDR < 0.05, alongside positive and negative control outcomes. Parentheses indicate the proxy exposure used for each gene. (c) Pathway-specific modulation of a shared MRI feature by distinct drug-target genes. (d) Comparison of cis-MR (MR-LD-IVW) and polygenic MR estimates for selected gene-MRI feature pairs; colored points denote cluster-specific polygenic MR estimates indicated by MR-Cluster. (e) MR scatter plot for LDL-C on CAD, with SNPs grouped into pathway-specific clusters by MR-Cluster; rs11591147 and rs12916 denote the lead variants for *PCSK9* and *HMGCR*, respectively. (f) Analogous plot for TG on CAD; rs964184 and rs11207980 denote the lead variants for *APOC3* and *ANGPTL3*, respectively.

We identified one MRI biomarker linked to *GLP1R (firstorder_Minimum_inp)* and two linked to *APOC3 (glszm_ZoneEntropy_inp* and *glszm_GrayLevelVariance_inp)*, with no pathway-linked features identified for *LPA, PCSK9*, or *HMGCR —* consistent with the weaker genetic correlations observed between Lp(a)/LDL-C and liver MRI features (Figure 5e). Negative control analyses yielded no unexpected findings, and estimation precision remained acceptable across methods (Figure 7b). The *APOC3*-linked feature *glszm_ZoneEntropy_inp* reflects spatial heterogeneity of hepatic in-phase signal intensity, and its association with *APOC3* aligns with the gene’s central role in TG metabolism, suggesting that impaired TG clearance may drive heterogeneous hepatic lipid deposition detectable by radiomic texture entropy. Notably, the cis-MR effect directions on *glszm_ZoneEntropy_inp* differed between *APOC3* and *HMGCR: APOC3*-mediated TG elevation was associated with decreased *glszm_ZoneEntropy_inp*, whereas *HMGCR*-mediated LDL-C elevation showed the opposite direction (MR-IVW est = 0.196, P = 0.106; robust MR methods P < 0.05; Figure 7c), suggesting that TG- and cholesterol-lowering pathways modulate hepatic signal heterogeneity through mechanistically distinct routes. This discrepancy is supported by convergent but independent lines of evidence. The *APOC3* direction aligns with MR estimates of *glszm_ZoneEntropy_inp* on downstream metabolic traits (cluster C2; Figure 6b), where *glszm_ZoneEntropy_inp* showed a negative effect on HbA1c and a positive effect on HDL-C, a pattern coherent with TG-related metabolic signaling. The *HMGCR* direction, in turn, is consistent with the positive genome-wide genetic correlation between *glszm_ZoneEntropy_inp* and LDL-C (cluster C7; Figure 5e), reflecting the broader landscape of cholesterol-related hepatic effects. For *GLP1R*, high heterogeneity in BMI instruments (Cochran’s Q P < 0.001) motivates us to use hypothalamic gene expression data as a sensitivity analysis, where we confirmed the significant *GLP1R* effects on both *firstorder_Minimum_inp* (est = -0.063, P = 0.022) and BMI (est = -0.558, P < 0.001). These findings collectively highlight the complex, pathway-dependent genetic architecture of liver radiomic features.

To validate that identified MRI features reflect gene-specific pathway effects rather than proxy exposure effects, we compared cis-MR estimates against polygenic MR estimates of the same proxy exposure on each highlighted MRI outcome; a divergence between the two would support the MRI feature as a genuine pathway-linked biomarker independent of the proxy trait (illustrated by Figure 7a). Since polygenic MR effects may cluster into distinct pathways, we account for this heterogeneity using MR-Cluster when comparing against cis-MR estimates (Methods). As an illustration, the cis-MR results for LDL-C on CAD differed between *PCSK9* (MR-LD-IVW = 0.527–0.694) and *HMGCR* (MR-LD-IVW = 0.241–0.516) (Figure 7d), and their corresponding lead variants (rs11591147 and rs12916) are located in two clusters indicated by MR-Cluster (Figure 7e). This likely reflects pleiotropic effects of statin-mimicking *HMGCR* variants on T2D and BMI [17–19], and the two clustered effects of PCSK9 and HMGCR are consistent with recent drug-target MR findings [20,21]. Polygenic MR showed no effect of BMI on *firstorder_Minimum_inp* (est = 0.009, P = 0.632), diverging significantly from the cis-MR estimate (heterogeneity P < 0.0001 ; Figure 7d). Our genetic correlation analysis similarly found no BMI–*firstorder_Minimum_inp* association (Figure 5e; Supplementary Table S25-S27). These results support *firstorder_Minimum_inp* as a possible valid biomarker of *GLP1R* pathway activity. Analogously, polygenic MR for TG on *glszm_GrayLevelVariance_inp* yielded null or positive clustered estimates (overall est = 0.020, P = 0.410), contrasting with the negative APOC3 cis-MR estimate (heterogeneity P = 0.0006; Figure 7d), suggesting that *glszm_GrayLevelVariance_inp* could capture an APOC3-specific biological pathway beyond general TG effects. For *glszm_ZoneEntropy_inp*, cis-MR via APOC3 and polygenic MR yielded divergent estimates for TG, but concordant estimates for LDL-C (Figure 7d).

These findings demonstrate that liver MRI features could capture gene pathway-specific effects diverging from conventional proxy-exposure MR estimates, potentially serving as mechanistically informative biomarkers for evaluating the downstream impact of targeted therapeutic interventions.

## Discussion

The genetic architecture of high-dimensional liver imaging phenotypes and their connections to external traits have remained largely unexplored. Here, we derive 200 well-defined liver MRI radiomic features using deep learning-based segmentation and radiomics extraction from nearly 43,000 UK Biobank participants, characterize their epidemiological and genetic architecture through population-scale association analyses and hypothesis-free GWAS, and investigate their biological relationships with external traits through multi-omics integration and a range of genetic structural and causal inference frameworks.

Two overarching findings emerge from this work. First, high-dimensional liver radiomic features capture heritable genetic information that is structurally distinct from, and largely orthogonal to, liver fat (PDFF), the most widely studied hepatic imaging phenotype [4,5]. Analyses of cross-locus pleiotropy profiles, latent genetic structure, and heritability enrichment converge on the same conclusion: the radiomic feature space encodes genuine hepatic biological variation beyond fat-correlated redundancy, including signals from loci with no prior association with liver fat and enrichment in liver-specific regulatory annotations. Second, these features are not passive imaging readouts but mechanistically informative biomarkers of specific biological pathways. Genetic correlations establish their broad cardiometabolic relevance at the genome-wide level; Mendelian randomization reveals directional causal connections with non-redundant pathway structure per outcome; and cis-MR demonstrates that individual features can capture gene-level pathway perturbations in ways that diverge from conventional proxy-exposure approaches. Moving from genome-wide overlap through causal inference to pathway anchoring, this analytical hierarchy collectively positions liver radiomic features as a new class of imaging biomarkers with mechanistic resolution beyond conventional endpoints.

Looking forward, three directions appear promising. First, the fat-independent associations identified here motivate clinical application of radiomic features in liver disease subtyping. Among MRI features significantly associated with incident chronic liver disease after full adjustment for liver fat and iron, texture features were prominent, suggesting these features capture fibrosis- or inflammation-related tissue remodeling rather than steatosis per se. Complementing these epidemiological findings, Mendelian randomization independently identified several water-based MRI features with consistent causal connections to liver-related outcomes, providing genetic-level evidence that changes in these imaging traits may reflect, and potentially precede, progressive hepatic pathology along the steatosis-to-cirrhosis spectrum. Of particular translational relevance, MASH drug development currently relies on liver biopsy for staging and treatment response assessment, and these converging epidemiological and causal findings raise the possibility that radiomic texture features could serve as non-invasive imaging biomarkers capable of tracking subclinical tissue changes across disease stages, potentially reducing reliance on biopsy in future therapeutic trials. Second, the divergent cross-feature effect profiles of genetic variants with distinct mechanisms, illustrated by the concordant *PNPLA3/TM6SF2* signature versus the more dispersed *APOE* profile across 59 MRI features, suggest that high-dimensional radiomic space could serve as a functional annotation layer for genetic variants. Just as transcriptomic or proteomic profiles have been used to characterize variant mechanisms, MRI feature profiles may enable data-driven clustering of variants by their hepatic phenotypic consequences, providing imaging-grounded insight into pathway specificity. Third, our Genomic SEM results caution against naive aggregation of radiomic features by imaging category, but simultaneously point toward a constructive solution: features loading onto shared latent factors may be legitimately combined in multi-trait frameworks to boost statistical power, while features loading onto orthogonal factors should be analyzed separately to preserve biological resolution. More broadly, the causal structures identified here, linking specific MRI features to distinct hepatic pathways, may inform the construction of decoupled or composite phenotypes [15,22] that more precisely index underlying hepatic biology, potentially improving both the power and interpretability of future investigations.

Several limitations merit consideration. Our GWAS cohort is predominantly of European ancestry, and while replication in the UKB non-European independent set yielded concordant effect directions for most variants, absolute replication rates were constrained by modest sample size. A more fundamental methodological tension concerns the choice between data-driven machine learning features and the well-defined radiomic features employed here: while end-to-end learned representations may offer superior predictive performance, well-defined radiomic features confer interpretability, reproducibility, and cross-study harmonizability essential for genetic analyses, a design tradeoff that future studies incorporating both paradigms could address. More broadly, the complex genetic architecture of high-dimensional liver MRI features poses a challenge permeating all post-GWAS analyses: genetic boundaries between features do not align with intuitive radiomic categories, correlations are pervasive and structured, and many post-GWAS frameworks may contend with partially overlapping genetic information. While we addressed this through complementary approaches, including genetic correlation clustering, exposure selection within MVMR, and cis-MR for pathway anchoring, studies working with lower-heritability features or complex genetic structures should exercise particular caution in interpreting genetic overlap results derived from high-dimensional imaging phenotypes.

In summary, high-dimensional liver MRI radiomic features derived from population-scale imaging encode heritable biological variation with a multi-factorial genetic architecture extending beyond hepatic fat, demonstrate widespread associations with circulating proteins, metabolites, and cardiometabolic traits, and connect to a range of disease outcomes through potentially distinct causal pathways. These findings could establish a genetic and epidemiological foundation for integrating liver radiomic imaging into the mechanistic investigation of common diseases and the evaluation of therapeutically relevant hepatic pathways.

## Method

### UK Biobank

The UK Biobank recruited more than 500,000 participants aged 40–69 years across the United Kingdom between 2006 and 2010, with extensive phenotypic and genetic data collected at baseline, including surveys, biospecimens, anthropometrics, vital signs, and other study-specific assessments. The UK Biobank imaging substudy aimed to acquire abdominal MRI scans in up to 100,000 participants aged 40–69 years. Approximately 50,000 participants underwent abdominal MRI as part of a reinvitation protocol commencing in 2014. Imaging was performed on 1.5 T clinical MRI scanners (Magnetom Aera, Siemens Healthineers, Erlangen, Germany) using whole-body T1-weighted dual-echo gradient echo (GRE) sequences with the following acquisition parameters: echo times of 2.39/4.77 ms, pixel size of 2.23 x 2.23 mm^2^, slice thickness of 3–4.5 mm, repetition time of 6.69 ms, and flip angle of 10°. Four imaging contrasts were acquired per participant: in-phase, out-of-phase, water, and fat. All analyses of UK Biobank data were conducted under UK Biobank application 7089.

### Extraction of liver radiomics features

Liver radiomic feature extraction comprised two main steps: (1) segmentation, in which liver voxels were identified from the full abdominal volume, and (2) feature extraction, in which radiomic features were computed from the segmented liver region. Liver segmentation was performed using a deep learning algorithm applied to abdominal MRI scans from 43,316 participants. Only samples for which liver segments could be successfully identified by our segmentation algorithm were retained for downstream quantitative analysis, yielding a final imaging cohort of 43,174 participants. Radiomic features were subsequently extracted using *PyRadiomics* (v3.1.0), yielding 200 liver radiomic features per individual as defined by the software’s feature library ([website](https://pyradiomics.readthedocs.io/en/latest/features.html)).

### Segmentation

Whole-body MRI scans were first reconstructed by stitching together six acquisition station blocks per individual using a stitching algorithm [23], yielding four Dixon contrast volumes per participant: water, fat, inphase, and out-of-phase. Participants missing any of the four contrast phases were excluded. Liver segmentation was performed using a pre-trained nnU-Net model [24], a self-configuring variant of the U-Net architecture that has demonstrated superior performance across a range of biomedical segmentation tasks. The model accepts all four Dixon contrasts as input channels and outputs a 3D voxel-wise annotation map, where a value of 1 indicates a liver voxel. The model was initially trained on 200 manually labelled abdominal MRI scans, each from the UK Biobank and GNCC cohorts (400 total, labelled by radiologists), with performance validated via 4-fold crossvalidation. It was subsequently applied to 20,000 scans (including 10,000 from the UK Biobank) and reviewed manually by radiologists, achieving an error-free segmentation rate exceeding 95%, so no additional fine-tuning was performed in the present study. Segmentation was conducted on Google Cloud Platform using Tesla T4 GPUs (16 GB VRAM, CUDA 11.3). The resulting liver segmentation mask was applied to the whole-body image to define the liver region of interest (ROI) for each participant, with each voxel assigned a gray-level intensity value across the four Dixon contrasts. Radiomic features were subsequently extracted from the water and inphase contrasts only. The fat and out-of-phase contrasts were excluded as they predominantly reflect hepatic lipid content, information largely captured by proton density fat fraction (PDFF), and thus may offer limited biological resolution beyond existing fat-based measures. Water-phase images were retained as they are sensitive to tissue hydration, inflammation, and fibrosis, while in-phase images integrate both water and fat compartments to characterize overall hepatocellular tissue signal intensity.

### Feature extraction

Radiomic features were computed from the liver ROI and mask using *PyRadiomics* (v3.1.0), applied separately to water and in-phase images. Each contrast yielded 107 features, referred to as water features and in-phase features respectively, for a total of 214 extracted features. Features were organized into three main categories. Shape features describe the 3D size and geometry of the liver based solely on the segmentation mask, independent of voxel intensity values; as these are contrast-independent, the 14 shape features were identical across water and in-phase images and were therefore deduplicated, retaining a single set. First-order features summarize the statistical distribution of voxel intensities within the liver ROI. Texture features quantify spatial relationships among neighboring voxels using matrix-based representations, including the gray level co-occurrence matrix (GLCM), gray level run length matrix (GLRLM), and gray level size zone matrix (GLSZM), among others. All extracted features are continuous phenotypes. After removing duplicate shape features, 200 valid radiomic features were retained for downstream analysis. Full definitions of all radiomic features are available at website.

### Epidemiological analysis

#### Quality control and principal components analysis

Due to the pervasive skewness observed across liver radiomic features, all features were winsorized at a threshold of five median absolute deviations (MAD) from the median prior to association analyses, affecting on average 1.843% of samples per feature. This procedure mitigates the influence of extreme values while preserving biological interpretability. Features with pairwise Pearson correlations |r| > 0.9 were iteratively pruned until all remaining pairs satisfied |r| ≤ 0.9, to reduce redundancy and avoid duplicated association signals in downstream analyses. Principal components analysis (PCA) was subsequently conducted on all retained MRI features both jointly and separately within each feature category (shape, first-order, and texture). To characterize the relative compactness of the phenotypic variance structure within each category, we defined the effective dimensionality proportion as the number of principal components required to explain 80% of the total variance divided by the total number of features in that category.

#### Phenotypic correlation analysis

We first examined the association between BMI at the MRI visit and each retained liver radiomic feature, fitting a linear regression of each feature on age, age^2^, sex, sexxage, sexxage^2^, and BMI. We then investigated phenotypic associations between liver radiomic features and a panel of cardiometabolic traits, including liver fat, liver iron, LDL-C, HDL-C, TG, Lp(a), HbA1c, and coronary artery disease (CAD). For each continuous trait, values exceeding five times the MAD from the median were excluded prior to analysis to mitigate outlier influence. Associations were estimated by regressing each liver MRI feature on the trait of interest, with adjustment for age, age2, sex, sexxage, sexxage2, and BMI. More details on the phenotypes, including quality control, visit instance, and medication adjustment, are provided in the Supplementary Note S1.

#### Proteome- and metabolome-wide association analyses

Using the same covariate-djusted regression framework, we extended phenotypic correlation analyses to 2,923 plasma proteins measured on the Olink Explore 3072 platform (UKB-PPP) and 325 NMR metabolites. For each protein or metabolite, we regressed each retained radiomic feature on the analyte level, adjusting for age, age^2^, sex, sexxage, sexxage^2^, and BMI, with outlier removal as described above. To assess the contribution of hepatic fat content to observed associations, all analyses were repeated with additional adjustment for liver PDFF (Field 21088). Significance was determined using Bonferroni correction across all analyte-radiomic pairs tested.

#### Incident disease analysis

We evaluated associations between liver radiomic features and three incident endpoints: coronary artery disease (CAD), type 2 diabetes (T2D), and chronic liver disease (CLD), with time zero defined as the imaging visit date and individuals with prevalent disease at the imaging visit were excluded. Associations were assessed using Cox proportional hazards models under three covariate adjustment strategies: minimal (age, age^2^, sex, agexsex, age^2^xsex, BMI), augmented (additionally including disease-specific risk factors), and full (further adjusted for liver fat and iron). Cox-LASSO with unpenalized clinical covariates was used to identify jointly independent radiomic predictors under the full model. Predictive performance was evaluated by 5-fold cross-validation, comparing augmented covariates alone, plus liver fat and iron, and plus LASSO-selected radiomic features, using time-dependent AUROC at 1, 3, and 5 years. Full details are provided in the Supplementary Note S1.

### Genome-wide association studies

#### Participant quality control

Participant-level quality control followed criteria established in recent imaging GWAS of comparable UK Biobank imaging cohorts [25]. To minimize confounding from population stratification, analyses were restricted to participants of self-reported White ancestry, with additional outlier exclusion based on genetic ancestry PCA using the R package *aberrant*, as described in prior studies [6,26]. Participants with individual-level SNP missingness exceeding 10% were also excluded. After applying these filters, 37,725 participants were retained for GWAS, with similar sample sizes reported in previous UK Biobank imaging GWAS of liver fat [6,5].

#### Phenotyping

All 200 liver radiomic features were quantitative phenotypes. Given the pronounced skewness observed across many features, rank-based inverse normal transformation (RINT; Blom’s transformation with c = 3/8 [27]) was applied to all features prior to association analyses to mitigate the adverse effects of non-normality on GWAS results [28]. RINT was applied before residualization, implemented via the *--apply-rint* flag in REGENIE. This approach effectively removes the influence of outlier values common in imaging phenotypes [29], enabling all 200 features to be retained for genetic association analyses. Residuals were then obtained by regressing each RINT-transformed feature on age, age^2^, sex, agexsex, age^2^xsex, genotyping array, and the first 10 ancestry principal components. These residuals were used as the phenotypes in all subsequent GWAS analyses and are referred to as MRI features throughout unless otherwise stated. We note that some prior imaging GWAS, including those for liver fat, performed residualization prior to RINT. While this ordering may be statistically justifiable [30], we found it unsuitable for our radiomic features, as it resulted in residual associations between the transformed phenotypes and covariates, and produced unreliable GWAS results for features with extreme outlier values.

#### Genotyping

UK Biobank genotyping procedures have been described in detail previously [31]. Participants were genotyped using either the UK BiLEVE Axiom Array or the UK Biobank Axiom Array (Affymetrix). Array-derived genotypes were imputed using a combined reference panel comprising the Haplotype Reference Consortium, UK10K, and 1000 Genomes Project.

#### Association analysis

Genome-wide association studies (GWAS) were conducted separately for each of the 200 liver radiomic features across chromosomes 1–22 using REGENIE (v3.2.8). Rather than excluding related individuals, sample relatedness was accounted for within the REGENIE framework. REGENIE employs a two-step fitting procedure. In Step 1, variants from the UK Biobank genotyping array were filtered using the following quality control criteria: minor allele frequency (MAF) < 1%, genotyping rate < 90%, and Hardy-Weinberg equilibrium P < 1×10^−15^. The retained variants were used to construct whole-genome genetic predictors for each individual via ridge regression, capturing polygenic background and accounting for sample relatedness. In Step 2, linear regression was fitted for each phenotype conditional on these genetic predictors. Imputed UK Biobank genotypes (version 3) were used as the test variants, restricted to those with MAF > 1% and imputation INFO score > 0.8, yielding 8,182,636 variants for association testing.

#### GWAS of BMI-adjusted MRI features

Prior liver MRI studies have examined adiposity-adjusted imaging phenotypes [4,32], and we here additionally conducted GWAS with BMI included as a covariate. We note that adjustment for heritable covariates such as BMI may introduce collider bias and yield misleading associations [33,34]. BMI-adjusted GWAS were conducted using the same procedures as the primary analysis, with the addition of BMI as a covariate. Two BMI measurements were considered: BMI at baseline (instance 0) and BMI at the imaging visit (instance 2). Although instance 2 BMI is more phenotypically correlated with liver MRI phenotypes, its use as a covariate could be more susceptible to over-adjustment and collider bias. The unadjusted GWAS (without BMI adjustment) was designated as the primary analysis, with BMI-adjusted results used for sensitivity and downstream genetic correlation analyses, considering the confounding role of obesity.

#### GWAS diagnostics

Genomic control inflation factors (λGC) were computed for each radiomic feature, and LD score (LDSC) regression was performed using LDSC (v1.0.0) with the HapMap3 SNP list to assess potential confounding from population stratification and cryptic relatedness. Summary statistics were processed using the munge function with the following exclusion criteria: MAF < 0.01, imputation INFO score < 0.9, strand-ambiguous variants, and variants within the major histocompatibility complex (MHC) region on chromosome 6. LDSC regression was then fitted using HapMap3 SNPs with MAF > 0.05. Features with an LDSC intercept exceeding 1.1 were considered to reflect substantial sample structure confounding and were excluded from all downstream analyses. The diagnostic results are given in Supplementary Table S4.

### Heritability and genetic correlation among radiomic features

SNP-heritability estimates and their standard errors were computed for all 200 liver radiomic features using univariate LDSC regression. Pairwise genetic correlations across all 19,900 feature pairs were estimated using cross-trait LDSC regression. To reduce genetic redundancy in downstream analyses, features were iteratively pruned such that no pair retained an absolute genetic correlation exceeding 0.9, yielding a final set of 59 liver MRI features for post-GWAS analyses.

### Functional enrichment analysis

Partitioned LD score regression (S-LDSC) was conducted using the most recent 97-annotation baseline model alongside cell-type-specific binary annotations. Baseline LD scores, cell-type annotations, variant weights, and allele frequency files based on the European 1000 Genomes Project Phase 3 were obtained from the Google Cloud Bucket ([website](https://console.cloud.google.com/storage/browser/broad-alkesgroup-public-requester-pays/LDSCORE)). Heritability enrichment, defined as the proportion of SNP-heritability attributable to a given annotation relative to the proportion of SNPs it contains, was estimated for liver-specific annotations across the 9 lead MRI features using the *--h2* flag.

### Risk locus definition

Because the MRI traits are highly and complexly correlated, applying a multiple-testing correction would make the genome-wide significance threshold overly conservative. We therefore used 5×10^−8^ as the suggestive threshold. For each liver MRI feature, approximately independent lead variants were identified using PLINK (v1.9) with 1000 Genomes Project European reference data. Variants with MAF > 0.01 were clumped using a 1 Mb window and an LD threshold of r^2^ < 0.01, retaining variants that reached genome-wide significance (P < 5×10^−8^). Each lead variant was annotated to the nearest protein-coding gene within ±0.1 Mb using the Ensembl database. When aggregating results across multiple MRI features, lead variants from all features were pooled, and any two lead variants located within 1 Mb of each other with rs^2^ > 0.2 were assigned to the same locus.

### Internal pleiotropy analysis

We assessed whether identified risk loci were associated with multiple liver MRI features, defining pleiotropy as a locus reaching genome-wide significance (P < 5×10^−8^) for more than one feature. Pleiotropic loci were visualized across the 10 lead liver MRI features, comprising the 9 new radiomic features with the highest SNP-heritability z-scores and liver fat (PDFF), using circular plots generated with *Circos* [35,36]. To characterize the broader phenotypic consequences of MRI-associated variants, their associations with other traits were queried from the NHGRI-EBI GWaS Catalog using the R package *gwasrapidd* (v 0.99.18; used at Mar 26, 2026).

### Replication analysis

In the absence of an independent external cohort with liver radiomic features, replication was first conducted using an internal split-sample strategy within the UK Biobank. The imaging dataset was divided into a discovery set (80%; n = 30,227) and a validation set (20%; n = 7,564), stratified by age and sex. Feature transformation and REGENIE were performed after the data split to ensure independence between subsets. Independent lead variants were identified in the discovery set by LD clumping at a genome-wide significance threshold (P < 5×10^−8^). Replication in the validation set was assessed using the same association model, with a variant considered replicated if it showed (i) a concordant direction of effect and (ii) nominal significance (P < 0.05). Internal replication results were only used to assess the robustness of GWAS findings; all effect estimates reported in downstream analyses were derived from the full discovery sample.

A second replication analysis was conducted in UK Biobank participants not included in the primary European ancestry GWAS, comprising ancestry PC-based White outliers (n = 2,658), Asian (n = 571), Black (n = 272), mixed ancestry (n = 191), and other ancestries (n = 203). Lead variants were defined based on the primary European GWAS at significance thresholds (P < 5×10 ^-8^), and directional concordance rates and nominal replication rates were evaluated in this multi-ancestry validation sample

### Genetic latent factor analysis

We used Genomic SEM to investigate the latent factor structure of liver MRI features based on their genetic covariance. Genomic SEM is a factor analysis framework in which the covariance matrix of the variables of interest is replaced by the genetic covariance matrix of the phenotypes, with additional model restrictions on the latent factors to learn the underlying factor structure. We obtained the LDSC-derived genetic correlation and covariance matrix of the liver MRI features and only focused on the 59 retained MRI features for the analysis to avoid potential non-invertibility of the matrix induced by high genetic correlations in downstream computations.

#### Genetic data preparation for Genomic SEM

Genetic covariance estimates were derived via LDSC regression, such that sample overlap across MRI features does not bias the estimates, as it is absorbed into the LDSC intercept term. For each radiomic feature category, the four most heritable features, defined by the z-score of SNP-heritability from univariate LDSC regression, were initially considered as candidates. Liver fat and liver cT1 were additionally included as established liver MRI phenotypes. Features were further filtered to retain only those with a heritability z-score exceeding 7, consistent with the threshold applied in the original S-LDSC paper [37], yielding 9 radiomic features plus liver fat (10 variables in total) for Genomic SEM analyses. This threshold was imposed because features with low statistical power for heritability estimation frequently introduced large errors during positive-definite matrix smoothing, rendering the Genomic SEM model fitting unreliable.

#### Genomic SEM fitting strategy

Genomic SEM analysis proceeded in two steps. In the first step, exploratory factor analysis (EFA) was conducted on the genetic covariance matrix to identify a putative loading structure. In the second step, confirmatory factor analysis (CFA) was used to formally specify and evaluate parametric factor models. To guard against overfitting, whereby latent factors directly reflect the observed genetic covariance matrix rather than underlying biological structure, multivariable LDSC regression was performed separately on even and odd chromosomes to obtain independent estimates of the genetic covariance matrix and its associated sampling covariance matrix (estimated via jackknife, using the *GenomicSEM* package). The even-chromosome genetic covariance matrix was used exclusively for EFA in Step 1, while the odd-chromosome estimates were reserved for CFA model fitting in Step 2. To validate this chromosome-splitting approach in our liver MRI analysis, QQ plots of z-statistics comparing odd- and even-chromosome genetic covariance estimates were examined and showed approximately linear behavior without evidence of tail inflation (Supplementary Figure S7), indicating that the two sets of estimates were statistically consistent within expected sampling variability.

#### Exploratory factor analysis

In Step 1, the genetic covariance matrix was first smoothed to the nearest positive definite matrix using the R function *nearPD*, and exploratory factor analysis was performed with the number of factors ranging from 1 to 5 using the *factanal* package in R. Both promax rotation (permitting correlated latent factors) and varimax rotation (enforcing orthogonal factors) were applied. For each solution, factor loadings with absolute values exceeding 0.3 were retained; for any feature with no loading meeting this threshold, the largest absolute loading was retained to ensure all features were represented. The loading structures identified across the 10 candidate factor solutions were carried forward to inform the confirmatory model specifications in Step 2. Factor solutions with more than five factors were not considered, as higher-dimensional models frequently resulted in convergence failures or poorly identified solutions, including non-positive-semidefinite latent factor covariance matrices. This is consistent with observations reported in recent work [38].

#### Confirmatory factor analysis

In Step 2, all 10 candidate factor models were fitted using the odd-chromosome genetic covariance matrix, and model fit was evaluated using AIC, CFI, and SRMR. Higher CFI values indicate better fit, and SRMR values below 0.10 are considered acceptable [12]. Where negative residual variances were encountered, a known occurrence in Genomic SEM fitting, residual variances were constrained to be positive (> 0.001) and the model was refitted prior to model comparison. Only models with SRMR < 0.10 were retained for consideration, and the best-fitting model was selected based on the lowest AIC and highest CFI. This was the five-factor model identified under promax rotation in the EFA step (odd-chromosome fit: AIC = 269.78, CFI = 0.997, SRMR = 0.056). The selected model was subsequently refitted using the full autosomal genetic covariance matrix, yielding similarly good fit metrics (AIC = 297.01, CFI = 0.998, SRMR = 0.052), and this final model was used for all main text results and downstream analyses. All results are presented on the standardized scale of genetic correlations among features and latent factors.

#### Hierarchical factor analysis

The moderate genetic correlations observed among the five latent factors motivated a further hierarchical factor analysis. Exploratory factor analysis was first conducted on the estimated genetic correlation matrix of the five latent factors using the *factanal* package in R with varimax rotation and two higher-order factors, yielding an orthogonal two-factor solution. This model is fully identified, with 10 estimable parameters corresponding to the 10 off-diagonal entries of the 5×5 latent factor correlation matrix. The resulting loadings and uniqueness parameters indicated that latent factors F1 and F4 loaded predominantly onto the first and second higher-order factors, respectively (Supplementary Table S16). A confirmatory hierarchical Genomic SEM model was subsequently fitted using this loading structure across all autosomes.

### External pleiotropy analysis

To better investigate the external pleiotropy of liver MRI feature-associated variants, we leveraged PLATLAS, a large-scale pleiotropy platform comprising >1,000 traits from major global biobanks (MVP, UKB, FinnGen, BBJ, ToMMo, and KoGES) with ancestry-stratified GWAS meta-analyses [39]. We obtained EUR-ancestry GWAMA summary statistics for 1,119 traits across 18 categories (Supplementary Table S10) for the 77 significant variants at 51 loci associated with the 9 lead MRI features. To avoid redundancy from LD within the same locus, one representative variant per locus was retained, yielding 51 variants for pleiotropy analysis. External pleiotropy was defined using a Bonferroni-corrected threshold of P < 0.05/(1,119 * 51). Based on prior genomic SEM analysis, the 9 lead MRI features were grouped into 4 genetic factors (F1, F2, F3, F5), which we adopted as the classification framework for variants as the internal pleiotropy pattern, as this better captures the underlying genetic architecture than MRI-derived categories. The 51 variants were accordingly categorized by their internal pleiotropy across the 4 factors (yielding 7 distinct patterns; Supplementary Figure S5) and characterized by their external pleiotropy across the 1,119 PLATLAS traits.

### Harmonization of GWAS summary statistics for external traits

Thirteen external traits were included in post-GWAS analyses, encompassing BMI [40], lipid traits [41,42], liver-related diseases [43,44], liver iron-corrected T1 (cT1; UkB Field 40062), type 2 diabetes (T2D) [45], and CAD [46]. For each trait, the most recent large-scale GWAS summary statistics were obtained from the NHGRI-EBI GWAS Catalog; full details are provided in the Supplementary Table S13. All external GWAS were restricted to European-ancestry populations with combined-sex analyses.

For each variant of interest (e.g., lead SNPs from our radiomic feature GWAS), association statistics, including effect size and standard error, were extracted from each external GWAS and harmonized to the same effect allele. All variants were checked for strand consistency across GWAS results to avoid misalignment of ambiguous palindromic SNPs (those with effect allele frequency among 0.45 and 0.55). For variants absent in the external GWAS, proxy variants were identified from the 1000 Genomes Project European reference panel as the highest-LD variant satisfying r^2^ > 0.8 with the original variant. Effect allele harmonization between the original and proxy variants was then performed using *LDlink* with the European population reference, and the proxy variant summary statistics were substituted for the original variant in all downstream analyses.

### Proteome-wide association study (PWAS) and colocalization

To identify plasma proteins associated with liver MRI features, we performed a PWAS using a two-sample two-stage least squares (2SLS) framework. Cis-pQTL information was obtained from the UKB-PPP study, which reports fine-mapped cis-pQTL results using strictly in-sample LD with SuSiE for all available proteins assayed on the Olink Explore platform [47]. In our analysis, we constructed two non-overlapping sets of unrelated European-ancestry UK Biobank participants. The MRI dataset (n=33,122) comprised unrelated European-ancestry participants with liver MRI data available, and the protein dataset (n=42,803) comprised the UKB-PPP participants, after excluding related individuals and the overlapping participants included in the MRI dataset. For each protein available in UKB-PPP on the autosomes (1916 proteins in total), we extracted dosages for the fine-mapped lead cis-pQTLs in all possible credible sets and estimated variant conditional weights in the first stage regression using the protein dataset by RINTed protein levels on cis-pQTL dosages and the covariate set as our GWAS. We then constructed a genetically predicted protein score for each individual in the MRI dataset using these weights from the first stage regression, and tested associations between genetically predicted protein levels and 59 RINT liver MRI features at the second stage using the same covariate adjustment. Significance was defined using a global Benjamini–Hochberg false discovery rate (FDR) < 0.05 across all tested protein-MRI pairs.

To assess whether PWAS-identified protein–MRI associations were driven by shared causal variants rather than linkage disequilibrium (LD) between distinct signals, we further performed colocalization analyses for all significant protein–MRI pairs. Full GWAS summary statistics for each protein were downloaded from the UKB-PPP Synapse repository (syn51365303). For each selected protein, the cis-region was defined as the span of all fine-mapped lead cis-pQTL positions, extended by ±500 kb. MRI GWAS summary statistics within the corresponding genomic window were extracted from our GWAS. All statistics from the protein and MRI traits were then harmonized and restricted to the shared variants for colocalization. We also constructed a nearly in-sample LD matrix using genotype dosages from our UKB MRI cohort and harmonized it. We conducted colocalization using the package *coloc* (v5.2.3) with default prior settings to obtain posterior probabilities (PP1-PP4) for hypotheses H0–H4, where H4 corresponds to a shared causal variant. To allow for multiple causal variants within a locus, we additionally performed fine-mapping using SuSiE [48] via *susieR* (v0.14.2) with default settings for the MRI GWAS and the UkB-PPP protein GWAS. We then conducted the SuSiE-based colocalization [49] to obtain the posterior probabilities for H4. The SuSiE-based colocalization was restricted to pairs with sufficiently reliable fine-mapping results (e.g., convergence of the protein SuSiE models, adequate association signal in the MRI trait). A posterior probability H4 (PP4) > 0.8 was considered evidence of colocalization.

### Metabolomic characterization of PWAS-prioritized loci

We focused on the 16 loci prioritized by the preceding PWAS and colocalization analyses and evaluated their associations with 249 metabolites using European-ancestry summary statistics from the recent UKB-NMR cohort (n = 434,646) [14]. For each locus, we selected a single representative lead cis-pQTL, defined as the variant with the largest log10(BF) in the corresponding fine-mapping results [47], and extracted its association statistics from the UKB-NMR GWAS [14]. Statistical significance was assessed using a Bonferroni-corrected threshold of 0.05/(16 × 249). For metabolites showing Bonferroni-significant association with the lead cis-pQTL at a given locus, we further performed colocalization analysis between the corresponding protein and metabolite using harmonized summary statistics across shared variants, applying the same locus definition, LD reference and colocalization framework as described above.

### Genetic correlation between liver radiomic features and external traits

Bivariate LDSC regression was conducted for all pairs of liver radiomic features and external traits using the *GenomicSEM* R package (v0.0.5). Genetic correlation, defined as the correlation between per-variant effect sizes across all reference variants under an infinitesimal model with joint Gaussian random effects, was estimated for each pair. This approach is robust to sample overlap between GWAS datasets, as overlapping samples are absorbed into the LDSC intercept. For GWAS summary statistics in which z-scores and P values had been adjusted for genomic inflation, original unadjusted P values were recovered prior to LDSC analysis. For binary traits derived from meta-analyzed logistic regression, cohort-specific effective sample sizes were often unavailable; SNP-specific effective sample sizes were therefore approximated from the standard errors of meta-analyzed effect estimates and in-sample effect allele frequencies, following established approaches [50,51].

### Mendelian randomization analysis

Extensive MR analyses were conducted using the 59 retained liver radiomic features as nominal exposures in a hypothesis-free framework. When all instrumental variables are valid, the MR scatter plot displaying per-variant genetic associations with the exposure against those with the outcome is usually expected to show consistent alignment along a line whose slope corresponds to the causal effect estimate. This property holds even when the exposure is a proxy for a true upstream exposure, provided that the instruments affect the proxy exposure and the outcome only through the upstream exposure. Evidence of a non-zero MR effect therefore supports the existence of a causal connection between the observed exposure and the outcome, though causal interpretation should remain cautious given the specific model scenario (illustrated by Figure 6a). Compared to genetic correlation, which reflects genome-wide genetic overlap between traits to reflect the mechanism, MR leverages a small set of trait-specific genome-wide significant SNPs as instrumental variables to infer the directional causal effect of one trait on another. Therefore, genetic correlation and MR results could be different.

#### MR analysis design

The primary aim of our MR analysis was to prioritize liver radiomic features with genetic evidence supporting causal connections to cardiometabolic and liver-related outcomes. Our MR framework comprised three components: (1) clustering of genetically similar MRI features based on shared instrumental variants, (2) radiomics-wide univariable MR (UVMR) treating each MRI feature individually as the exposure, (3) and multivariable MR (MVMR) jointly modelling highlighted features to provide structural causal insights. All MR analyses used GWAS results from unadjusted MRI features to avoid potentially misleading signals introduced by heritable-covariate adjustment in GWAS. The rationale for this framework is grounded in the causal scenarios illustrated in Figure 6a. When a liver MRI feature (e.g., X2) has a direct causal effect on the outcome (Scenarios 1 & 2), UVMR will detect this signal provided core instrumental variable assumptions are satisfied. Genetically similar features (e.g., X1 in Scenario 2) sharing instruments with X2 may also yield UVMR signals even in the absence of a direct causal effect, due to genetic overlap. In the more realistic setting where MRI features are influenced by multiple latent hepatic factors (Scenario 3), UVMR can still detect signals; a feature that cleanly reflects a single latent factor (X1) will tend to yield a clearer signal than one influenced by multiple latent factors (X2), which may exhibit greater heterogeneity across instruments. These distinctions are reflected in MR scatter plots of per-variant exposure–outcome associations, which provide richer diagnostic information than summary numerical estimates alone and have been used extensively in recent liver MRI analyses [5]. Expected MR scatter plot patterns under each scenario are provided in the Supplementary Figure S15.

#### Genetic similarity clustering

We define genetically similar features in the MR context as those exhibiting proportional genetic associations across shared instrumental variants. Such features tend to produce similar UVMR signals when used as exposures. Notably, genetic similarity in this MR-specific sense is not equivalent to genome-wide genetic correlation estimated via bivariate LDSC [52], as MR relies only on a small set of strongly associated variants rather than the genome-wide signal. We therefore propose a MR-specific similarity metric based on the Cochran’s Q statistic [53,54], quantifying the consistency of genetic associations across instrumental variants between pairs of mRi features, and applied hierarchical clustering to the resulting Q-statistic-based similarity matrix. Clustering was performed using all independent genetic instruments across the 59 retained MRI features, yielding 10 clusters. Full methodological details are provided in the Supplementary Note S2.

#### Two-sample summary data MR

All MR analyses were conducted using GWAS summary statistics. For most external traits, the GWAS sample sizes substantially exceeded our imaging GWAS (e.g., >1,000,000 for LDL-C, HDL-C, and TG), resulting in minimal sample overlap and a roughly two-sample MR design. The exception was liver fat, which was derived from the same UK Biobank participants as our radiomic feature GWAS with a comparable sample size, resulting in substantial sample overlap.

#### Genetic instrument selection

For each of the 59 radiomic features, genetic instruments were selected as SNPs associated at P < 1×10^−6^, clumped to independence at r^2^ < 0.01. Variants additionally associated with BMI (P < 5×10^−4^) were excluded to reduce potential confounding through adiposity pathways. To test for invalid instruments arising from reverse causation, Steiger filtering [55] was applied to all selected instruments. For each SNP, a onesided Bonferroni-corrected test was used to identify variants explaining significantly more variance in the outcome than in the exposure; such variants were flagged as potentially invalid instruments and removed from downstream MR analyses, though exclusion of such SNPs is not always strictly necessary [56]. Effect alleles were then harmonized across exposure and outcome GWAS.

#### Sensitivity analysis adjusting for BMI

As a sensitivity analysis, BMI was additionally included as a covariate in a multivariable (bivariable) MR model for each radiomic feature to obtain the exposure-specific effect estimate, which was interpreted as the direct effect of the exposure on the outcome independent of BMI. MVMR was performed only when sufficient instruments were available for identification (i.e., ≥ 2 instruments).

#### MR estimation

The primary MR estimator was the random-effects inverse-variance weighted (IVW) method, or the Wald ratio when only a single instrument was available. As instrument selection and exposure association estimation were performed in the same sample, the winner’s curse may lead to modest underestimation of effect sizes. However, since our MR analyses were primarily conducted to test the sharp causal null hypothesis, and genetic association estimates with the outcome are only minimally affected by the degree of sample overlap in our setting, we expect the impact of winner’s curse on testing conclusions to be negligible [57,58].

#### Weak instrument bias, heterogeneity testing, and robust MR methods

Given the use of a relaxed significance threshold (P < 1×10^−6^) for instrument selection, the joint F-statistic was recorded for each exposure to assess weak instrument bias. A low joint F-statistic (conventionally < 10) indicates potential weak instrument problems, which inflate type I error in one-sample MR settings and reduce power in two-sample settings, the predominant design in our analyses. When multiple instruments were available, Cochran’s Q statistic was computed to assess instrument heterogeneity. Weighted-median, weighted-mode, and MR-RAPS were additionally applied as robust sensitivity analyses.

#### Feature selection criteria

Liver radiomic features were highlighted as putative causal exposures if they satisfied all of the following criteria: (i) Benjamini-Hochberg-adjusted UVMR-IVW (random effects) P < 0.05; (ii) at least 3 valid instruments remaining after exclusion of BMI-associated variants, enabling sensitivity analyses across variants; and (iii) a majority of robust MR methods (weighted-median, weighted-mode, MR-RAPS) yielding P < 0.05. Highlighted features were subsequently excluded if the MVMR effect estimate for that feature showed a statistically significant (P < 0.05) direction reversal relative to the UVMR estimate. All pairs with a joint F-statistic below 10 under the one-sample or overlapping case are removed. If the joint F statistic was below 10 in a strict two-sample setting, we iteratively removed the weakest instrument (lowest individual-variant F, computed as the squared t-value) one at a time (still retaining a minimum of 3) until joint F ≥ 10, and updated the MR results accordingly if feasible. Exposure-outcome pairs were not excluded solely on the basis of instrument heterogeneity (e.g., Cochran’s Q test) in our analysis, as heterogeneity is expected for our MRI features under certain plausible causal scenarios (e.g., Scenario 3; Figure 6a). MR calculations were performed using the R packages *MendelianRandomization* (v0.10.0) and *mr*.*raps* (v0.4.2).

#### Multivariable MR for the key MRI feature

Multiple significant UVMR signals for a given outcome do not necessarily indicate distinct causal pathways, as genetically similar MRI features, as reflected by our similarity clustering, may serve as surrogates for the same underlying latent factor. MVMR, which models multiple MRI features jointly as exposures, provides a more principled approach to disentangling correlated causal signals and is additionally more robust to horizontal pleiotropy mediated through other MRI features compared with UVMR. For each outcome, all radiomic features passing the UVMR selection criteria were included as candidate exposures in MVMR. Genetic instruments were defined as variants associated with any of the included features at P < 1×10^−6^, excluding BMI-associated variants, and clumped to independence at r^2^ < 0.01. Summary statistics for all clumped variants were collected for each radiomic feature exposure and each outcome, with effect alleles harmonized across datasets. Proxy variants from the European reference panel were used where direct variant matches were unavailable. For variable selection among correlated exposures, we applied MVMR-SuSiE [59,60], which incorporates the sum of single effects (SuSiE) framework into the MVMR model and performs soft variable selection on the exposures in the IVW regression while accounting for correlated instrument-exposure associations across exposures. MVMR-SuSiE is particularly suitable for highly correlated exposures such as our MRI features, and has been validated and previously applied in metabolomic MR contexts [60]. The number of key MRI latent factors for each outcome was indicated by the SuSiE algorithm, and the specific MRI features for each latent factor were those with larger posterior inclusion probabilities by MVMR-SuSiE. As a sensitivity analysis using a different model framework, MVMR-FA [61] was additionally conducted to investigate the latent causal structure among correlated MRI features using a factor analysis framework. More details of MVMR-SuSiE and MVMR-FA are given in Supplementary Note S2.

### Drug-target genes cis-MR analysis

Cis-MR analyses were conducted using genetic instruments restricted to the six established cardiometabolic drug-target genes — *GLP1R, LPA, PCSK9, HMGCR, APOC3*, and *ANGPTL3 —* to investigate the downstream effects of each gene pathway on the 59 liver MRI features in a hypothesis-free framework.

#### Data preparation

For each drug-target gene, plasma protein GWAS results were obtained from the UK Biobank Pharma Proteomics Project (UKB-PPP), and tissue-specific gene expression data were obtained from GTEx. In addition, established downstream proxy traits were selected for each gene based on known biological mechanisms and prior literature [62,63]: BMI for *GLP1R* (representing the obesity pathway), LDL-C for *HMGCR* and *PCSK9*, TG for *APOC3* and *ANGPTL3*, and Lp(a) for *LPA*. Brain hypothalamus tissue was used as the expression reference for *GLP1R*, given its relevance to the obesity pathway, while liver tissue was selected for all other genes. We note that the primary aim of the cis-MR analyses is to evaluate the effect of each drug-target gene on liver MRI features; cis-MR using proxy traits therefore remains valid provided the instruments satisfy core IV assumptions, though the causal interpretation differs between expression- or protein-based and proxy-traitbased cis-MR [64]. For *GLP1R*, the original BMI GWAS summary statistics [40] include a high proportion of UK Biobank participants, and weak instrument bias (F-statistic < 10) was observed for some cis-MR methods, rendering one-sample MR results unreliable due to inflated type I error. We therefore used BMI GWAS summary statistics derived from European participants in the MVP cohort [43] and refitted the cis-MR analyses for *GLP1R* to ensure a two-sample MR design.

#### Variant quality control

For each drug-target gene, common variants (MAF > 0.01) within a ±100 kb cis-window were considered as candidate instruments. Genetic instruments were selected by LD clumping (r^2^ < 0.01, P < 1×10^−6^) applied across all available exposure datasets (gene expression, plasma protein levels, and proxy traits). The exposure dataset yielding the greatest number of independent lead variants was selected as the primary exposure for downstream cis-MR estimation, maximizing instrument count to improve statistical power and enable robust sensitivity analyses. Although robust MR methods offer limited advantages in cis-MR settings due to the restricted number of available instruments [65], they can nonetheless better aid in identifying pleiotropic variants at certain loci (e.g., *GLP1R)*.

#### MR methods incorporating correlated variants

Compared with polygenic MR, cis-MR is less susceptible to horizontal pleiotropy due to its restricted genomic window [66], but is often underpowered owing to the limited number of independent association signals within any given cis-region. This limitation is particularly relevant in our hypothesis-free setting, where 59 liver MRI features are tested as outcomes. We therefore employed two complementary cis-MR methods that leverage correlated variants within the cis-region to do sensitivity analysis and increase statistical power beyond that achievable with conventional independent-instrument MR [67]. The first method, MR-LD-IVW, uses LD-based pruning to select variants iteratively, beginning with the most significant variant and adding variants whose pairwise r^2^ with all previously selected variants falls below a specified LD threshold, based on a reference LD matrix. Generalized IVW incorporating the LD matrix is then applied to obtain MR estimates. The second method, MR-PC-GMM, applies PCA to a weighted version of the LD matrix to derive the genetic principal components as instruments, with MR estimates consequently obtained via a generalized method of moments (GMM) framework. MR-LD-IVW has been shown to achieve the highest power in cis-MR analyses [67], while MR-PC-GMM offers greater robustness to numerical instabilities arising from misspecified or mismatched LD matrices [68,69–71], and the use of principal components as instruments is less sensitive to the choice of included SNPs. We also conduct the conventional MR analysis based on independent lead SNPs via random-effect IVW estimation (MR-IVW). Details on MV-LD-IVW and MR-PC-GMM are given in Supplementary Note S3.

#### Sensitivity analyses, positive and negative controls

Robust MR sensitivity analyses were conducted using weighted-median, weighted-mode, and MR-RAPS. As a positive control, CAD was tested as an outcome for all six drug-target genes. For *GLP1R*, cis-MR did not yield significant effects on CAD; examination of the MR-LD-IVW scatter plot suggested complex ratio estimates on CAD among variants (Supplementary Figure S16), likely due to the heterogeneous effects of *GLP1R* variants on multiple risk factors including BMI and HbA1c, as reported in recent studies [72]. T2D was therefore used as the positive control outcome for *GLP1R*, via its established obesity pathway. For all six genes, tanning ability (UKB Field ID 1727; obtained from the Neale Lab website) was used as a negative control outcome, following prior recommendations [73], to assess the estimation properties of the different cis-MR methods to identify any over-precise or unreliable estimates in our setting with (usually not perfect in-sample) LD information.

#### Cis-MR feature selection criteria

Liver MRI features were identified as putative drug-target pathway-linked biomarkers for a given gene if they satisfied all of the following criteria: (i) at least one of the three primary cis-MR methods (MR-IVW, MR-LD-IVW, or MR-PC-GMM) yielded an FDR-adjusted P < 0.05; (ii) all three primary cis-MR methods showed concordant effect directions with P < 0.05; and (iii) all robust sensitivity MR methods (weighted-median, weighted-mode, and MR-RAPS) showed concordant effect directions with P < 0.05.

#### Polygenic MR

For each proxy exposure used in cis-MR (BMI, LDL-C, and TG), polygenic MR was conducted using genome-wide significant instruments to estimate the exposure’s pervasive pathway-averaged effect on each selected liver MRI feature. A significant divergence between the cis-MR and polygenic MR estimates could indicate that the gene pathway exerts a direct effect on the MRI outcome independent of the proxy exposure, supporting the MRI feature as a valid pathway-specific biomarker. For BMI, 82 instruments were selected by LD clumping of GIANT summary statistics (P < 5×10^−8^, r^2^ < 0.01, MAF > 0.01), with genetic association estimates obtained from the same BMI dataset used in cis-MR (MVP cohort) to avoid winner’s curse bias. For LDL-C and TG, the winner’s curse is considered negligible given the large sample sizes of the respective GWAS [74]; the 100 most significant variants were therefore selected from the same GWAS used in cis-MR, using a more stringent clumping threshold (P < 5×10^−10^, r^2^ < 0.01, MAF > 0.01). LDL-C and TG instruments were distributed across multiple distinct loci, ensuring broad representation of polygenic pathway backgrounds. All instruments were harmonized to the effect allele of the MRI feature GWAS prior to MR estimation.

#### Variant clustering and cluster-specific MR analysis

To account for potential heterogeneity in polygenic MR estimates arising from distinct biological pathways, MR-Cluster [75] was applied to identify variant clusters with homogeneous per-variant causal effect estimates. MR-Cluster models the Wald ratio estimate of each variant as arising from a mixture distribution, where latent cluster membership with distinct effect sizes could reflect different underlying pathways. The conditional probability of cluster membership is estimated for each variant, and variants are assigned to the cluster with the highest posterior probability. A designated junk cluster captures outlier variants whose per-SNP Wald ratio estimates are inconsistent with any identified pathway cluster. Randomeffects IVW was conducted within each cluster to obtain cluster-specific MR estimates, and an overall polygenic MR estimate was derived using all non-junk variants. Cluster-specific estimates were compared against cis-MR estimates to assess pathway concordance. We note that cluster-specific polygenic MR estimates may be overly precise, as they are derived post hoc from variants selected to have similar per-SNP MR effects by construction. The heterogeneity comparison between cis-MR and polygenic MR cluster-specific estimates was therefore treated as heuristic.

## Data availability

Full summary statistics for the liver MRI features will be publicly available on the GWAS Catalog upon publication. The individual data for UKB is available upon application at http://www.ukbiobank.ac.uk. The individual data of UKB basic, imaging, proteomics, and metabolomics data used in our analysis were downloaded under the UK Biobank application number 7089. The summary statistics of UKB-PPP and UKB-NMR are publicly available and were downloaded from Synapse (Project ID: syn51365303) and GWAS Catalog (GCST90497049-GCST90501086). The summary statistics for gene expression can be accessed at the GTEx Portal. The summary statistics for other traits involved in our analysis are publicly available at consortium platforms (DIAGRAM and GIANT), the GWAS Catalog, and the Neale Lab site. The necessary data for LDSC-based analysis can be accessed at https://data.broadinstitute.org/alkesgroup/LDSCORE/. The PLATLAS data can be accessed at https://platlas.cels.anl.gov/.

## Code availability

The relevant code and scripts for our analyses will be available at https://github.com/HDTian/liverMRI. Publicly available tools used in this work include nnU-Net v2 (https://github.com/MIC-DKFZ/nnUNet), PyRadiomics (v3.1.0; https://pyradiomics.readthedocs.io/), REGENIE (v3.2.8; https://rgcgithub.github.io/regenie/), PLINK (v1.9; https://www.cog-genomics.org/plink/), Circos (https://circos.ca/), LDSC (v1.0.0; https://github.com/bulik/LDSC), GenomicSEM (v0.0.5; https://github.com/GenomicSEM/GenomicSEM), PLATLAS (v1.0.0; https://github.com/Verma-Lab/platlas), Ensembl (https://www.ensembl.org/), gwasrapidd (v0.99.18; https://github.com/ramiromagno/gwasrapidd), LDpair (https://ldlink.nih.gov/ldpair), dbSNP (https://www.ncbi.nlm.nih.gov/snp/), SuSiE/susieR (v0.14.2; https://stephenslab.github.io/susieR/), coloc (v5.2.3; https://github.com/chr1swallace/coloc), LocusZoom (https://locuszoom.org/), and MendelianRandomization (v0.10.0; CRAN R package). Detailed information on R and Python packages used in the analysis is provided in the corresponding scripts.

